# Optimal Control of Chlamydia Model with Vaccination

**DOI:** 10.1101/2020.09.09.20191072

**Authors:** U. B. Odionyenma, A. Omame, N.O. Ukanwoke, I. Nometa

## Abstract

This paper presents an SVEIRT epidemiological model in the human population with Chlamydia trachomatis. The model incorporated the vaccination class and investigated the role played by some control strategies in the dynamics of the disease (Chlamydia tracomatis). The reproduction number which helps in determining the rate of spread of the disease, was calculated using the method proosed by van den Driessche and Watmough. The local and global stability of the equlibrium points where established, where it was observed that the model is locally asymptotically stable if the reproduction number is less than unity, and globally stable if a certain threshold value is greater than unity or the re-nfection rate is zero. The effect of the re-infection rate on the global stability suggests the exhibition of the phenomenon of backward bifurcation of the model. The backward bifurcation of the system was later studied, and it shows that backward bifurcation will occur if the value of the bifurcation parameter ‘a’ is positive. The optimal control of the model shows the effect of different strategies in the transmission dynamicsof the disease and the cost effectivenes of each control pair. It was observed that the treatment and control effort gives the most cost effective combinations and at the same time the highest rate of disease avertion when compared to other stratagies. Sensitivity analysis of the parameters as shown in model, shows parameters that have high impact on the chosen classes.

## 1 INTRODUCTION

Mathematical models of Infectious diseases are normallly used to study the transmission dynamics, where knowledge of its epidemiology is used to predict what happens as time goes on, and also investigate the impact of interventions. Such analyses helps to inform policy decisions, especially where there are limited empirical data available for comparative evaluation of different interventions [9].

Chlamydia infection is seen as one of the most common sexually transmitted disease (STD) in humans worldwide especially in European countries [12] and the United States [13, 14] caused by the bacterium Chlamydia trachomatis. It was estimated that around 92 million Chlamydia infections occurred worldwide in 1999, including 50 million women and 42 million men [19]. Any sexually active person can be infected with Chlamydia. The risk of infection varies mostly on the age of the population [17]. It is observed that people in the 20–24 years age group are at much higher risk of being infected as on average. In 2010, the 20–24 years age group represented 36% of all reported cases which is the highest proportion among all age groups. Untreated chlamydia infection can result in pelvic inflammatory disease (PID) in women, which can lead to ectopic pregnancy and tubal factor infertility. Chlamydia can cause epididymitis in men [9, 10].

Matheatical models of diseases has been carried by many researchers [34, 35, 36, 5, 6] on different disease with different goals and objectives. The modelling and analysis of Chlamydia have been done by many researchers (see,[6, 5, 17, 9, 19, 14, 10]) where they studied the transmission dynamics of chlamydia from different view points and orientations; Sharma and Samanta [5] considered two treatment controls, which gives an optimal control problem relative to the epidemic model so as to minimize the exposed and infective populations as well as to minimize the cost of treatment.The basic reproduction number (*R*_0_) is calculated using the next-generation matrix method. The stability analysis of the model shows that the system is locally asymptotically stable at the disease-free equilibrium (DFE) *E*_0_ when *R*_0_ < 1. A work on new two-group deterministic model for Chlamydia trachomatis was designed and analyzed by Sharomi and Gumel[6] to gain insights into its transmission dynamics, where the model is shown to exhibit the phenomenon of backward bifurcation, where a stable disease-free equilibrium (DFE) co-exists with one or more stable endemic equilibria when the associated reproduction number is less than unity. The basic model is extended to incorporate the use of treatment for infectious individuals (including those who show disease symptoms and those who do not). Bifurcation analysis of some models was also carried in ([11, 8, 8, 27, 28]), which predicts parameters that causes the re-emergence of the disease. The method presented by Castillo-Chavez [23] have been widely used in the the investigation of this phenomenon.

Optimal control strategies of models can also be seeen in the following works [3, 7, 31, 4, 32, 33, 34, 35, 37]. This helps to investigate combined control measures and interventions that can help suggest steps in minimizing the impact of infectious diseases, where The Pontryagins Maximum Principle was used extensively in this sense. We have identified that till now no study has been done in investigating the transmission dynamics of Chlamydia tracomatis that incoporates vaccination as a class, with interest on the optimal control methods and costeffectiveness analysis of the applied control strategies. In this work, we assess the impact of vaccination, prevention effort, treatment effort and control against re-infection in control and management of Chlamydia tracomatis. To the best of our knowledge this has not been investigated before.

**Table 1:**
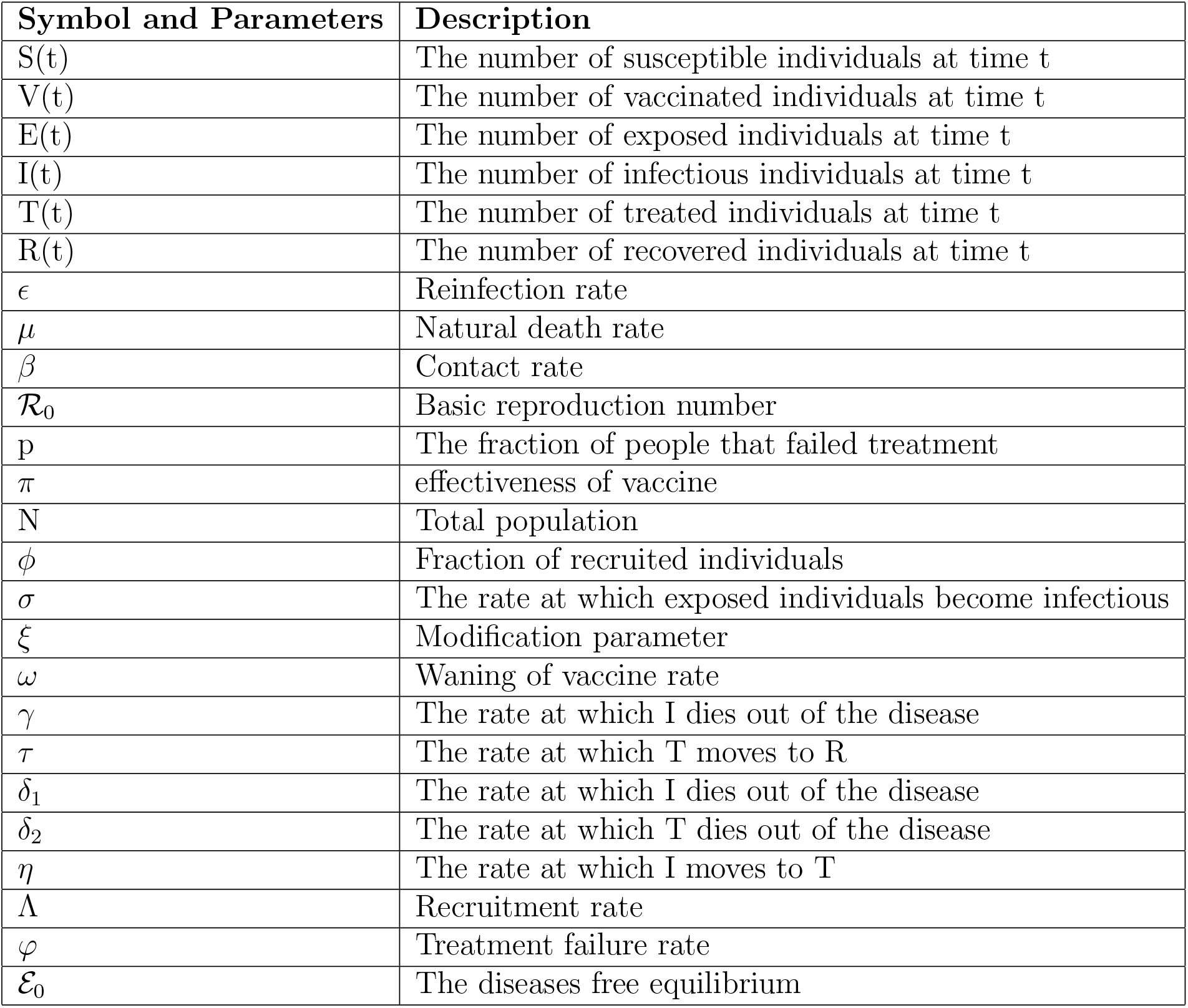
Parameter values and description

### 1.1 Symbols and Parameter Description Used in the Model

## 2 Mathematical Model Formulation

The total active population at time *t*, denoted by *N* (*t*) is divided into six compartments namely, the total susceptible population *S*(*t*), the total vaccinated population *V* (*t*), the total population of the exposed individual *E*(*t*), the total infected individual *I*(*t*), the total treated individuals *T* (*t*) and the total recovered individuals *R*(*t*) respectively.

*β* is the effective contact rate and Λ is the rate at which new individual are introduced into the susceptible population whereby a certain fraction is recruited into the vaccinated compartment. As time goes on the vaccine wanes out at rate *ω* and causes an increase in the susceptible population. Due to natural death, the population is reduced at the rate *µ*. The population of the infectious class is increased through the contact with the infected class and treated class. Also *ξ* is the modification parameter accounting for the reduced infectiousness of treated individuals compared to infectious individuals. Where the description of the other parameters are given in (Table 1).

Thus we have the model depicted below,

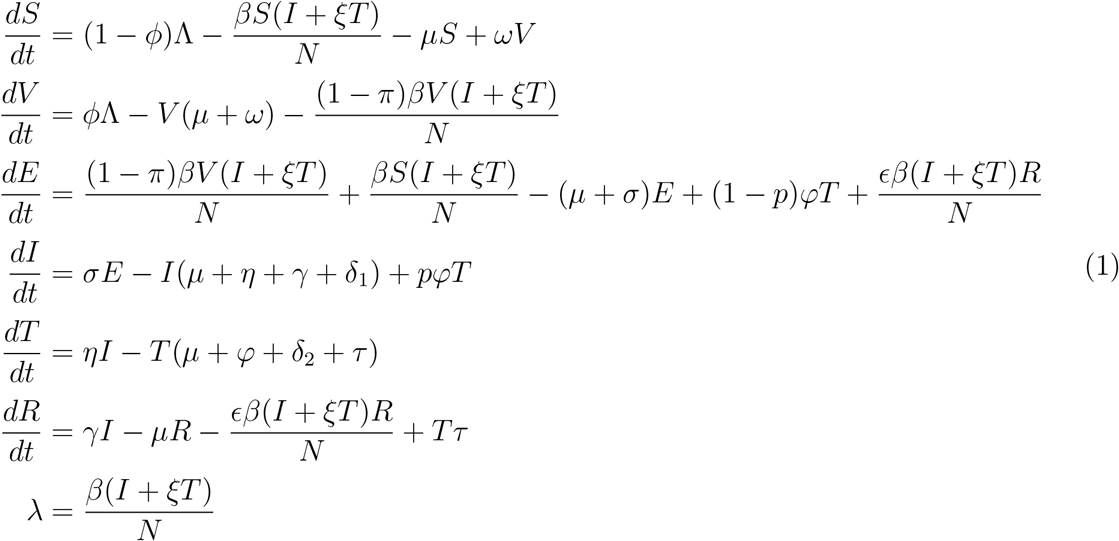

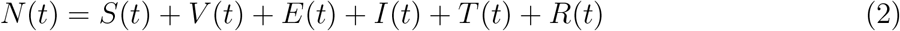

## 3 Basic Properties of The Model

### 3.1 Boundedness and Positivity of Solutions

The model will be analyzed in a biologically feasible region as follows. We first show that the system is dissipative (that is, all feasible solutions are uniformly-bounded) in a proper subset 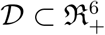

**Theorem 3.1**. *Consider the closed set* 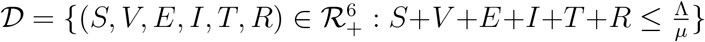 *is positively invariant*.

*Proof*. Adding all the equations of the model

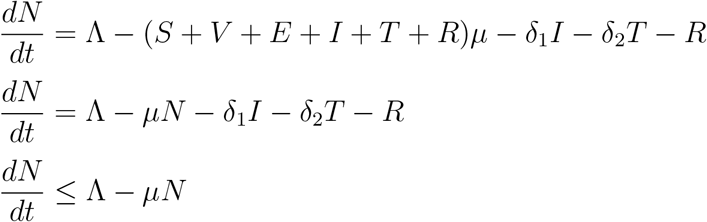

**Figure 1:**
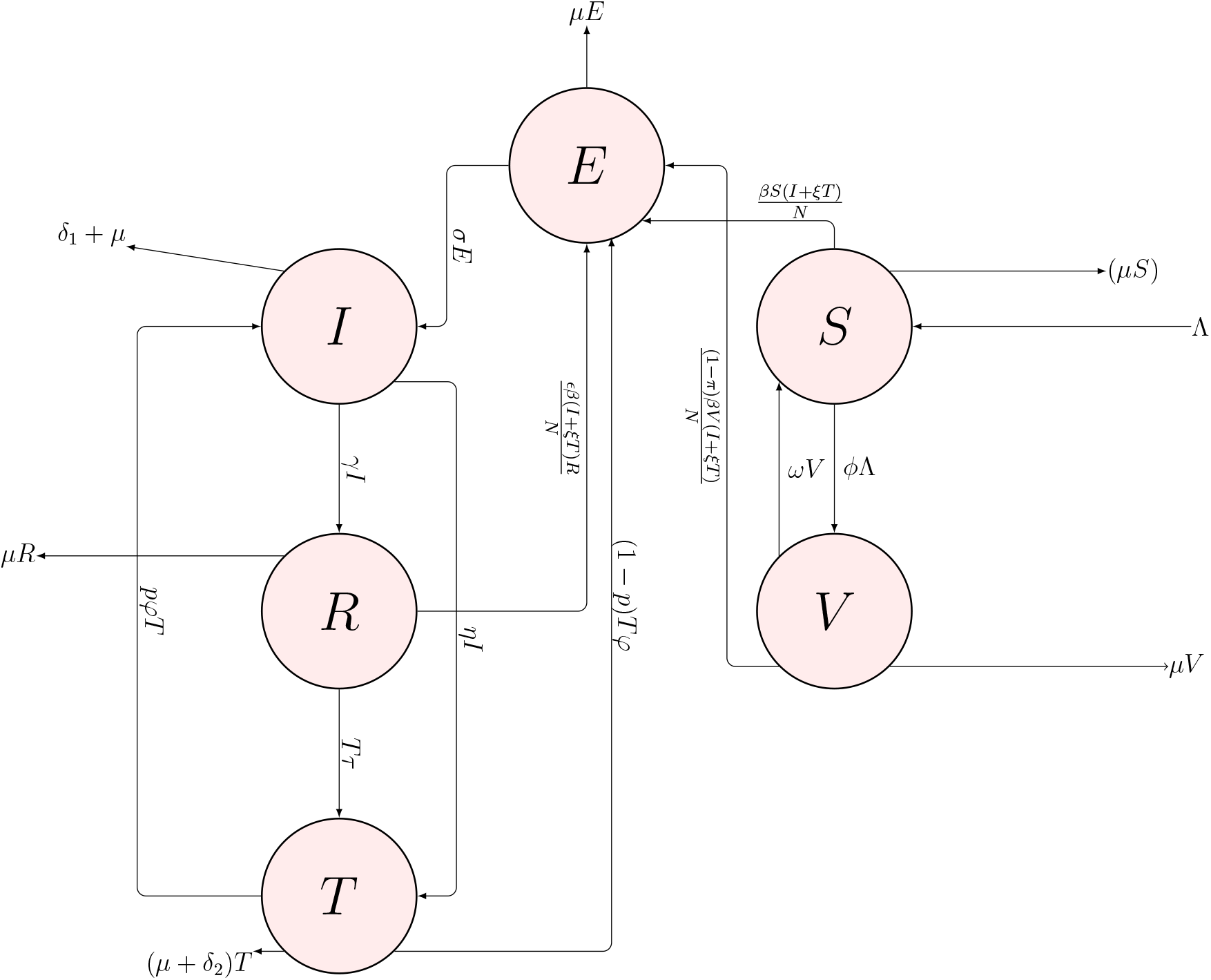
MODEL DIAGRAM

It follows that 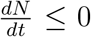 0 if 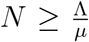. Thus a standard comparison theorem can be used to show that 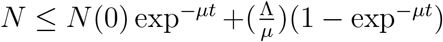. In particular, 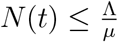 if 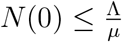.

Thus, the region 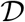 is positively invariant. Also, if 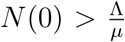, then either the solution enters 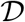 infinite time or N(t) approaches 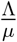 asymptotically.

Hence, the region 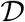 attracts all solutions in 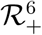

Since the region 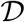 is positively invariant, it is sufficient to consider the dynamics of the flow generated by the model (1) in 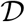, where the usual existence, uniqueness, continuation results hold for the system.

For the model equation to be epidemiologically meaningful, it is important to prove that all its state variables are non-negative for all time. In other words, solutions of the model system with positive initial data will remain positive for all time *t >* 0.

**Theorem 3.2**. *Let the initial data be S*(0) > 0*, V* (0) > 0*, E*(0) > 0*, I*(0) > 0*, T* (0) *>, R*(0) > 0*, Then the solutions S*(*t*)*, V* (*t*)*, E*(*t*)*, I*(*t*)*, T* (*t*)*, R*(*t*)) *of the model are positive for all time t >* 0.

*Proof*.

Let *t*1 = sup{*S*(*t*) > 0*, V* (*t*) > 0*, E*(*t*) > 0*, I*(*t*) > 0*, T* (*t*) > 0*, R*(*t*) > 0,}

Thus, *t*1 > 0.

It follows, from the second equation of (1) given as,

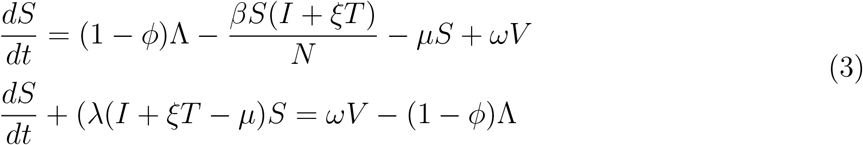

(3) is a linear differential equation, hence we shall solve by finding the integrating factor which is given as;

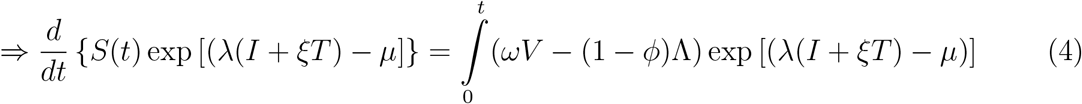

Now, if we integrate both sides of (4) from 0 to *t*_1_, we shall obtain:

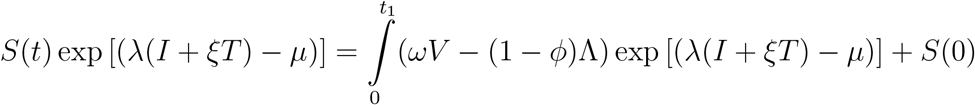

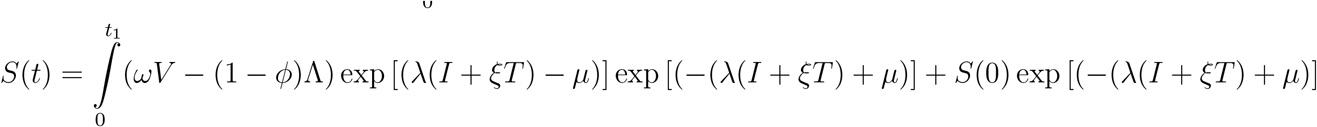

Hence, we shall observe that at *t*_1_ = 0, S(0) is always positive since exponential of a number is always positive. ⇒ *S*(*t*) > *S*(*t*_1_) > *S*(0). Since 0 is the sup of *S*(0) > 0. Then, S(t) is always positive. Similarly, the same proof shows that: *V* (*t*) > 0*, E*(*t*) > 0*, I*(*t*) > 0*, T* (*t*) > 0*, R*(*t*) > 0. which concludes that *S*(*t*)*, V* (*t*)*, E*(*t*)*, I*(*t*)*, T* (*t*)*, R*(*t*)) of the model are all positive for all time *t* > 0.

## 4 Analysis of The Model Without Control

In this section the formulated model will be analysed. We wiil calculate the basic reproduction number of the model and we also later investigated the existence and asymptotic stability of the steady (equilibrium) states.

### 4.1 Basic Reproduction Number ℛ_0_

The basic reproduction 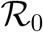 number is the average number of secondary infection that occur if a single infected individual is introduced into an entirely susceptible population.

The basic reproduction number 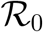 can also be defined as the effective number of secondary infections caused by an infected individual during his/her entire period of infectiousness [20]. This definition is given for the models that represent the spread of infection in a population.

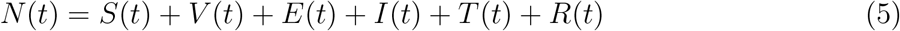

The basic reproduction number 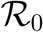 is given by taking the highest eigen value of the spectral radius *FV*^−1^ ([20]).

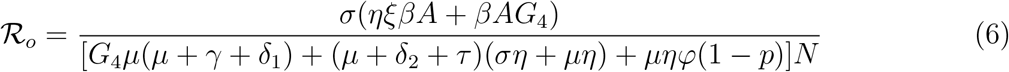

where, *A* = (*S*^∗^ + (1 − *π*)*V*^∗^)*, G*_1_ = *µ* + *ω, G*_2_ = *µ* + *σ, G*_3_ = *µ* + *η* = *γ* + *δ*_1_*, G*_4_ = *µ* + *ϕ* + *δ*_2_ + *τ N* = *S*^∗^ + *V*^∗^ + *E*^∗^ + *I*^∗^ + *T*^∗^ + *R*^∗^

### 4.2 Local Stability of the disease free equilibrium

The local stability of DFE(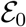) is established by using the next generation operator method on the model equation (1)

**Lemma 4.1**. *Using Theorem (2) in [20], we established that; For the model equation (1), the disease free equilibrium is locally asymptotically stable if the basic reproduction number* 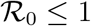 *and unstable if* 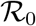 *is greater than unity (i.e*., 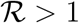).

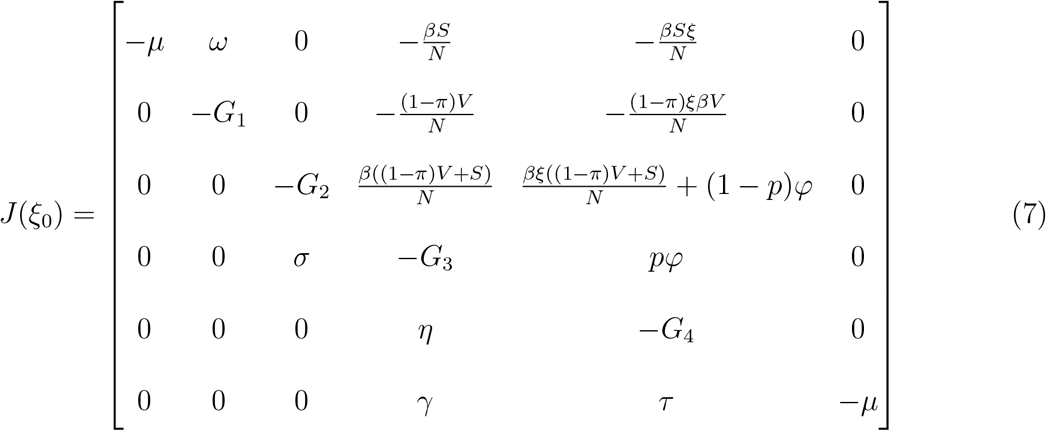

The characteristic equation is given as:

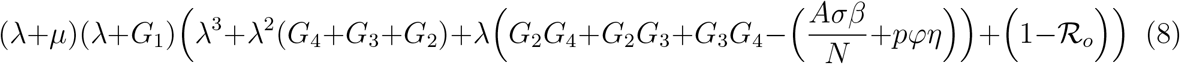

Using the Routh-Hurwitz criterion, the above polynomial (8) will have roots with negative real part if and only if 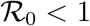.

#### 4.2.1 Endemic Equilibria of The Model

Steady state solutions are the solutions of the system of equations (1) when the right-hand side of a nonlinear system is set to zero. At the steady state, the model equation (1) is set to zero. That is, 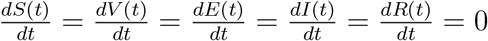

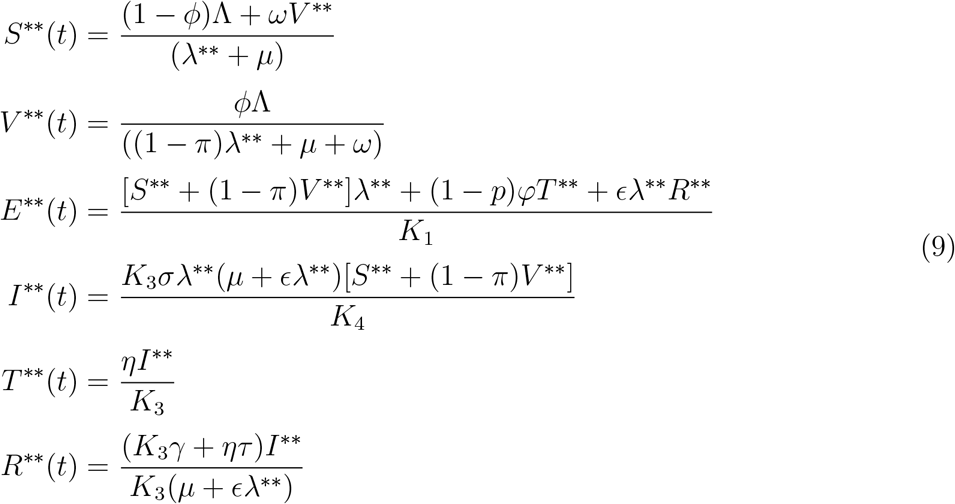

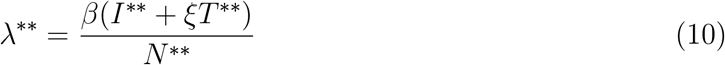

where, 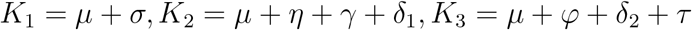 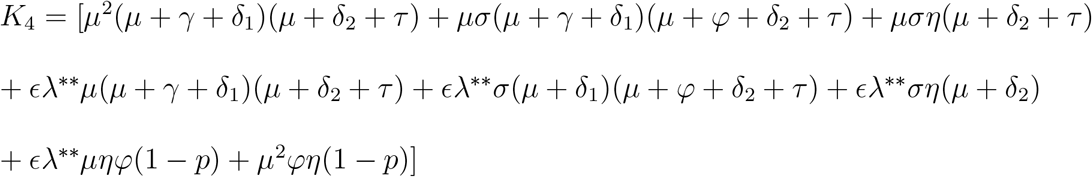

Hence from (10) *S*^∗∗^ + *V*^∗∗^ + *E*^∗∗^ + *T*^∗∗^ + *R*^∗∗^ = *βI*^∗∗^ + *βξT*^∗∗^

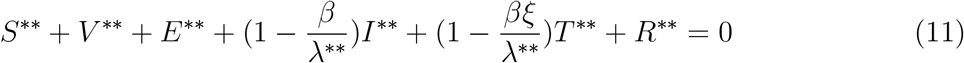

The endemic equilibria of (1) corresponds to the positive solutions of (11) above

### 4.3 Disease Free Equilibrium(DFE) *Ɛ*_0_

To determine the stability of the model, we first evaluate the equilibrium points or steady states of the ordinary differential equations (1). Steady state solutions or equilibrium points are the roots or solutions of the system of equations when the right-hand side of a nonlinear system is set to zero. The equilibrium point under consideration in this model is the Disease-Free (E = I = T = 0) Equilibrium point. At the disease free equilibrium, the disease compartments *E*^∗^ = *I*^∗^ = *T*^∗^ = 0

Hence the disease free equilibrium of (1) is given as;

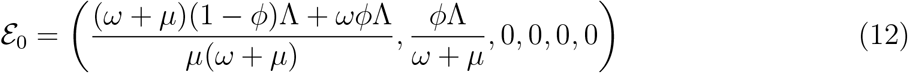

### 4.4 Global Asymptotic Stability(GAS) of the disease-free equilibrium(DFE) *Ɛ*_0_

. In this section, we list two conditions that if met, also guarantee the global asymptotic stability of the disease-free state. First, System (1) must be written in the form:

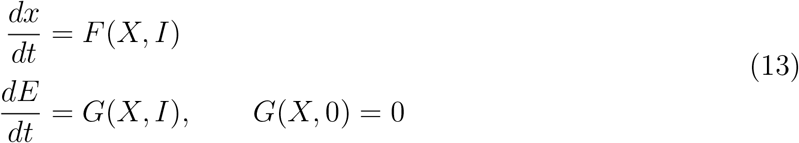

where *x* ∈ *R^m^* denotes (its components) the number of uninfected individuals and *I* ∈ *R^n^* denotes (its components) the number of infected individuals including latent, infectious, etc. *U_o_* = (*x*^∗^, 0) denotes the disease-free equilibrium of this system.

The conditions (H1) and (H2) below must be met to guarantee local asymptotic stability.

*dt*

(H1) For 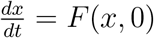, *x*^∗^ is globally asymptotically stable (g.a.s.),

(H2) 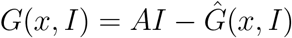, *G*(*x, I*) ≥ 0 for (*x, I*) ∈ Ω
where *A* = *D*_1_*G*(*x*^∗^, 0) is an M-matrix (the off diagonal elements of A are nonnegative) and *n* is the region where the model makes biological sense. If system (1) satisfies the above two conditions then the following theorem holds

**Theorem 4.1**. *Consider the model equation (1) with the DFE (12) given by* 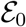, the *DFE* 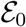 *of the model equation (1) is Globally Asymptotically Stable in* 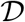 *whenever* 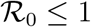.

We use the method presented in [23] to investigate the global stability.

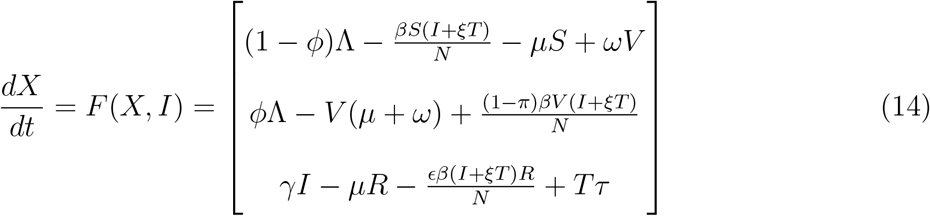

where ‘X’ denotes the number of non-infectious individuals and ‘I’ denotes the number of infected individuals

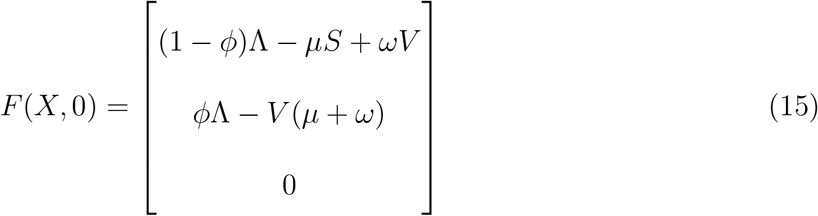

where ‘X’ denotes the number of non-infectious compartments and ‘I’ denotes the number of infectious compartments

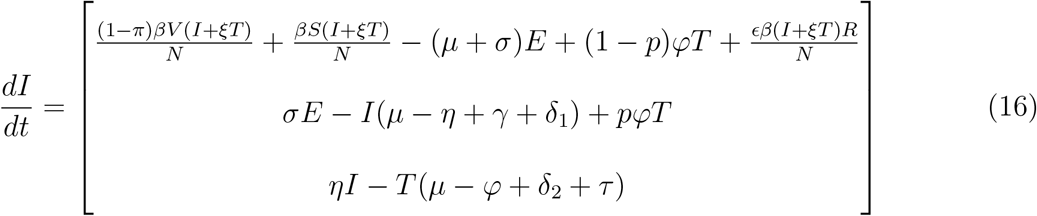

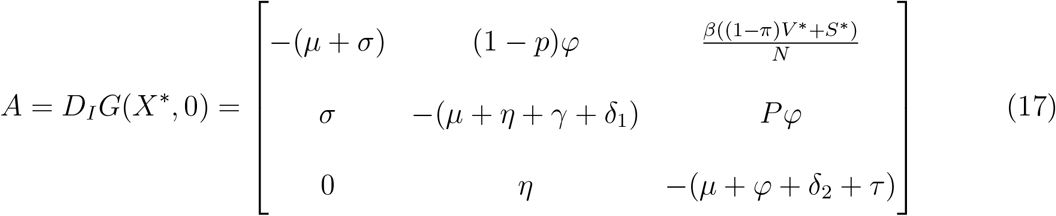

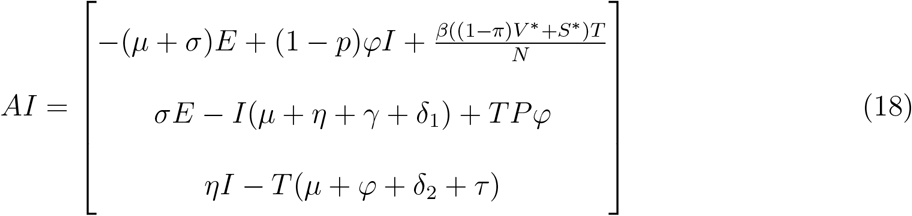

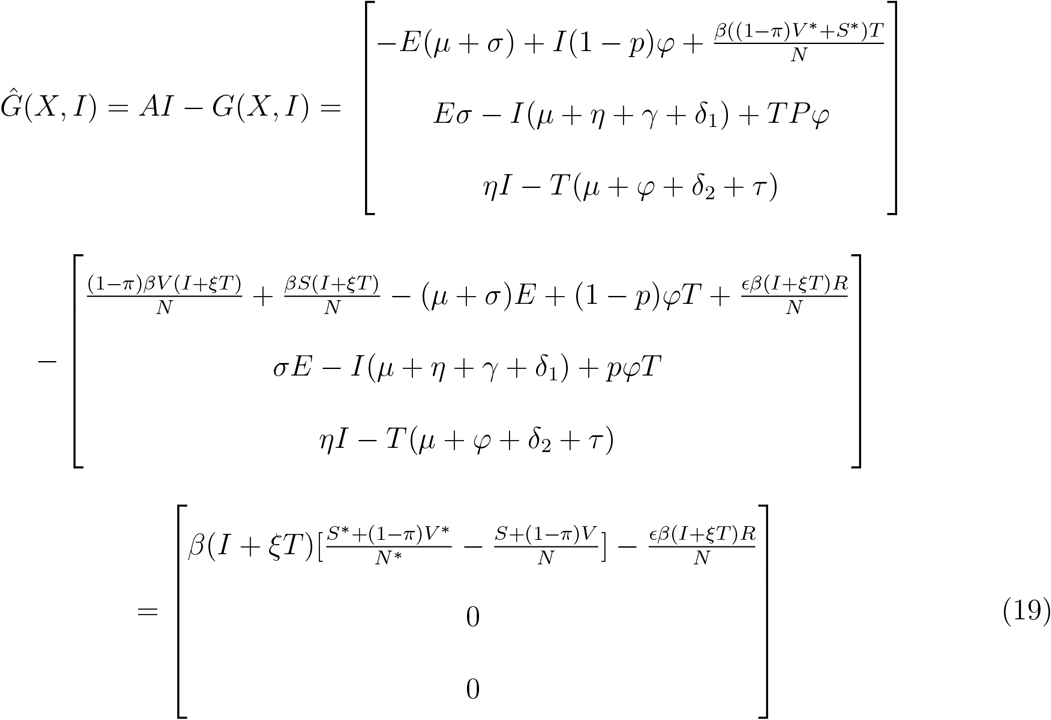

Since

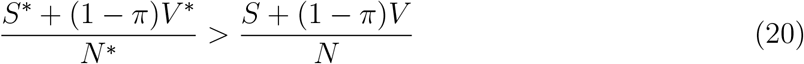

This implies 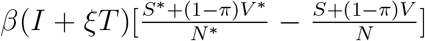 is positive, hence for the Disese Free Equilibrium(DFE) to be Globally Asymptotic Stable (GAS) either *ϵ* = 0 or 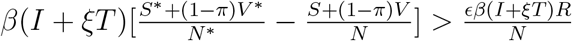

This shows the *ϵ* is a bifurcation parameter, hence this supports the backward bifurcation of the model. From figure 2 it shows that the re-infection rate which is suspected to be the cause of backward bifurcation, reduces the population of the recovered when increased, where this in line with the conclusion drawn above.

**Figure 2:**
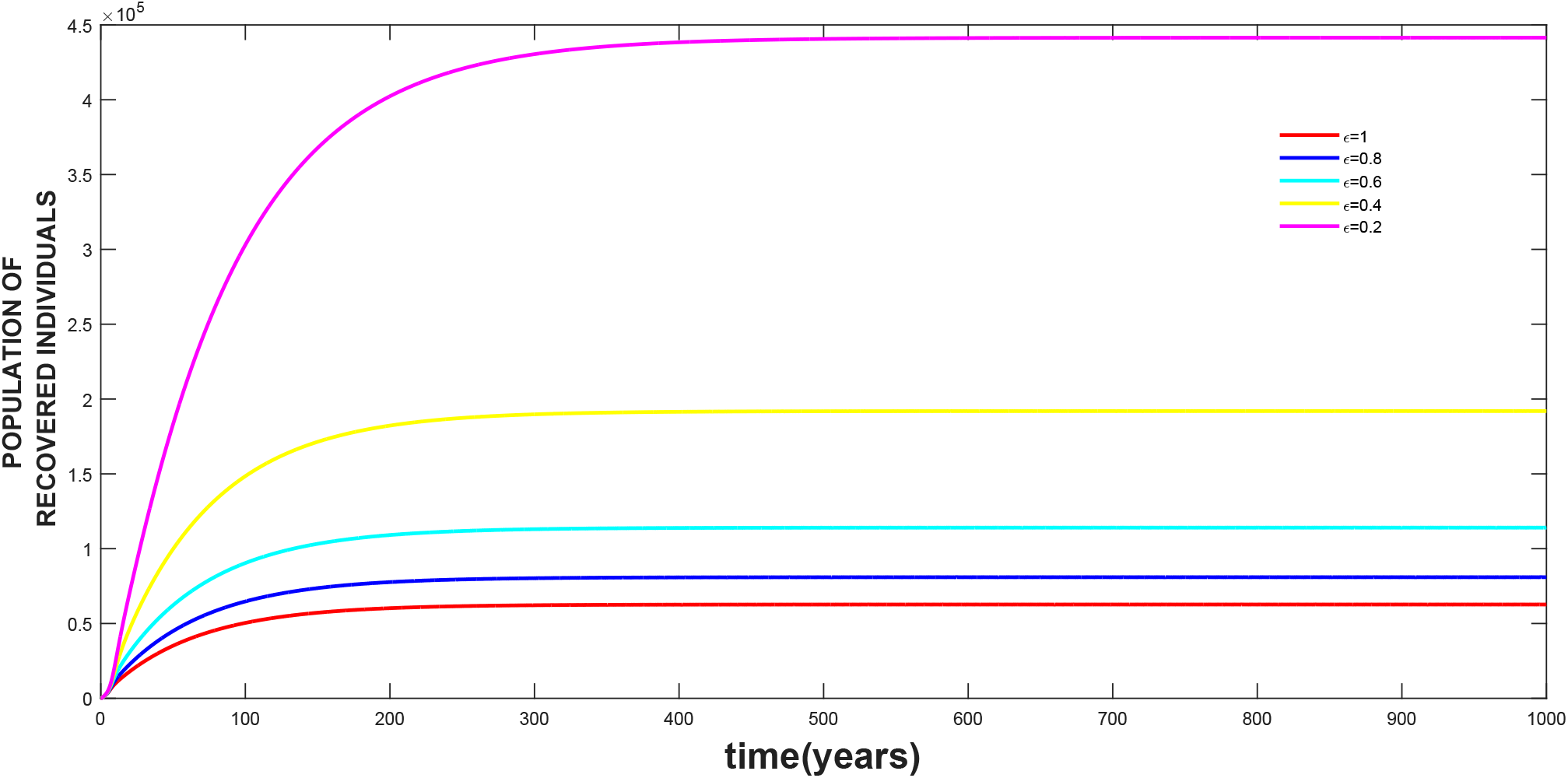
The effect of re-infection rate *ϵ* on the treated class, where the values of the other parameters are as given in table 3.

### 4.5 Bifurcation Analysis

In this section we investigate whether changes in some parmeters will result to a shift in the stability of the solution of the model. Using the Center Manifold Theorem as in [1], the following result is established.

**Theorem 4.2**. *Suppose a backward bifurcation coeffcient a >* 0 *(with ‘a’ defined below), when*

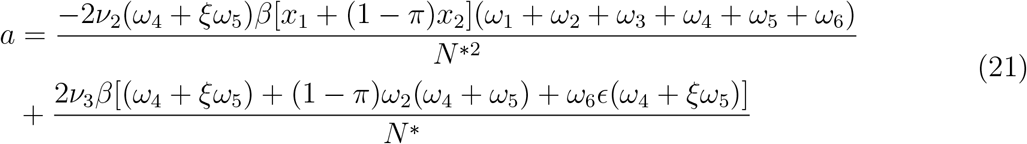

*then model 1 exihibits backward bifurcation at R*_0_ = 1*. If a <* 0*, then the system 1 exhibits a forward bifurcation at R*_0_ = 1

**Proof** Suppose *ξ_d_* = (*S*^∗∗^*, V*^∗∗^*, E*^∗∗^*, I*^∗∗^*, T*^∗∗^*, R*^∗∗^) represents arbitrary endemic equilibrium of the model, the existences of backward bifurcation will be studied using the Center Manifold Theory [1]. To apply this theory we will make the following substitutions;

let *S* = *x*_1_*, V* = *x*_2_*, E* = *x*_3_*, I* = *x*_4_*, T* = *x*_5_*, R* = *x*_6_,

so that 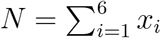

Further, using vector notation

*X* = (*x*_1_*, x*_2_*, x*_3_*, x*_4_*, x*_5_*, x*_6_)^*T*^

The model can be re-written in the form

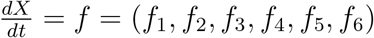 as follows;

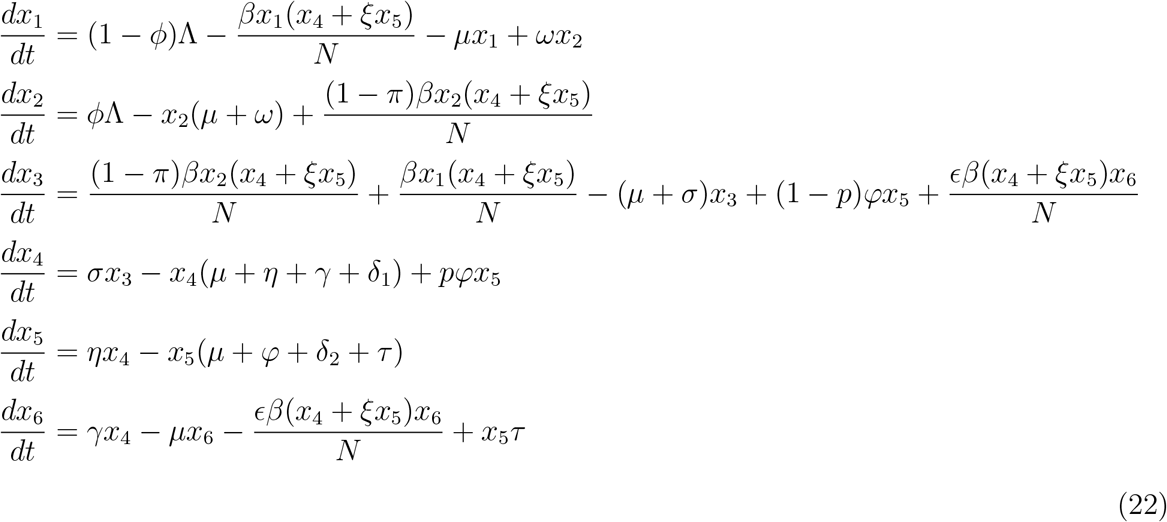

Finding the Jacobian of the model (22) at DFE, and using it to get the right eigen vectors is given below: Let

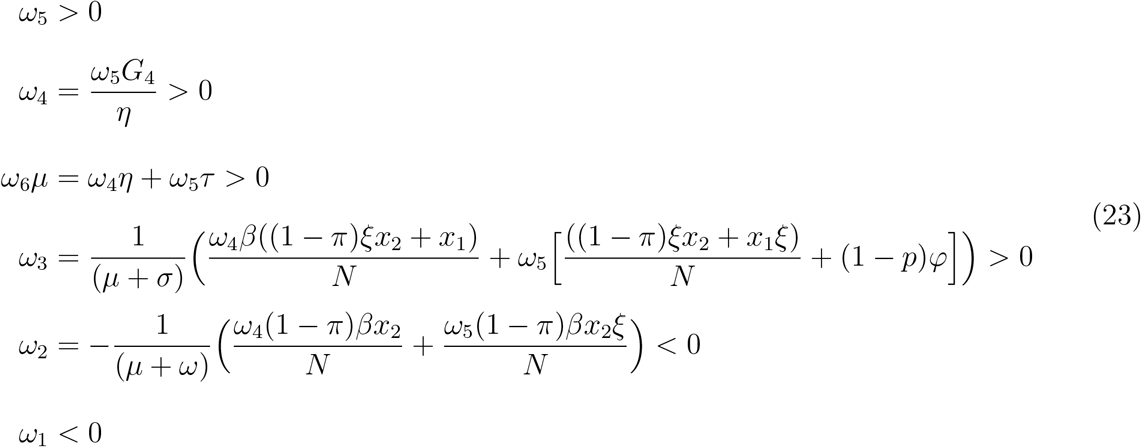

Calculating the left eigen vectors for the values of *ν*_1_*, ν*_2_*, ν*_3_*, ν*_4_*, ν*_5_*, ν*_6_ satisfyimg *ν* · *ω* = 1

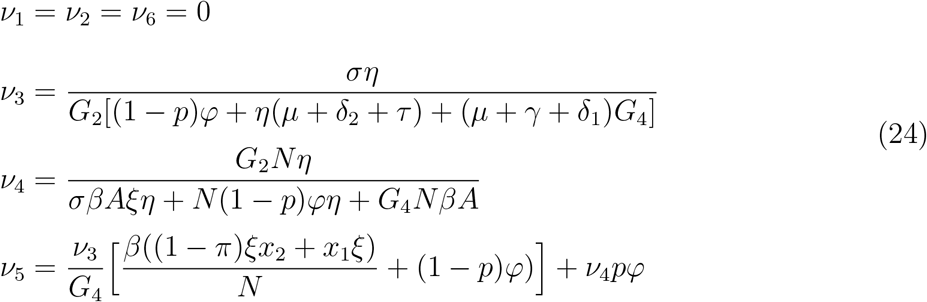

*ν*_3_ > 0*, ν*_4_ > 0*, ν*_5_ > 0 since all parameters are positive and (1 − *p*) > 0 then from

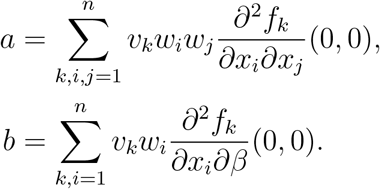

The local dynamics of the system around ‘0’ is totally determined by the sign of a and b.

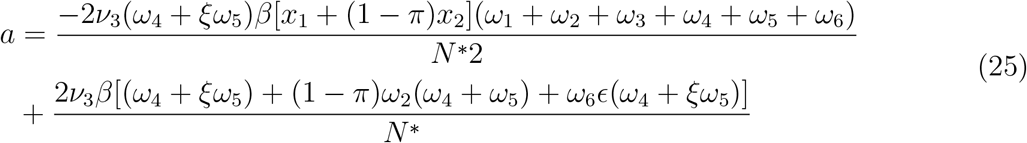

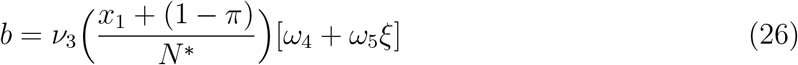

Since the bifurcation coefficient ‘b’ is positive, it follows from theorem 4.1 in [1] that the model (27), or the transformed model (22), will undergo backward bifurcation if the bifurcation coefficient ‘a’ given is positive i.e,;

i. if *a >* 0 and *b >* 0. When *β <* 0 with |*β*| 1, 0 is locally asymptotically stable and there exists a positive unstable equilibrium; when 0 ≤ *β* 1, 0 is unstable and there exists a negative, locally asymptotically stable equilibrium;
ii. If *a <* 0 and *b >* 0. When *β* changes from negative to positive, 0 changes its stability from stable to unstable. Correspondingly a negative unstable equilibrium becomes positive and locally asymptotically stable.

## 5 Optimal Control Analysis

In this section, the optimal control theory is applied to the system of differential equation modelling the population dynamics of Chlamydia. We shall use the Pontryagin’s Maximum Principle to determine the control measures that can be put in place to reduce the impact of Chlamydia tricomatis on the population. The controls include vaccination of a fraction of the poplation, prevention effort, control against re-infection and treatment effort against Chlamydia. The control model is solved numerically using the fourth order Runge-kutta method.

The optimaal control model is given as:

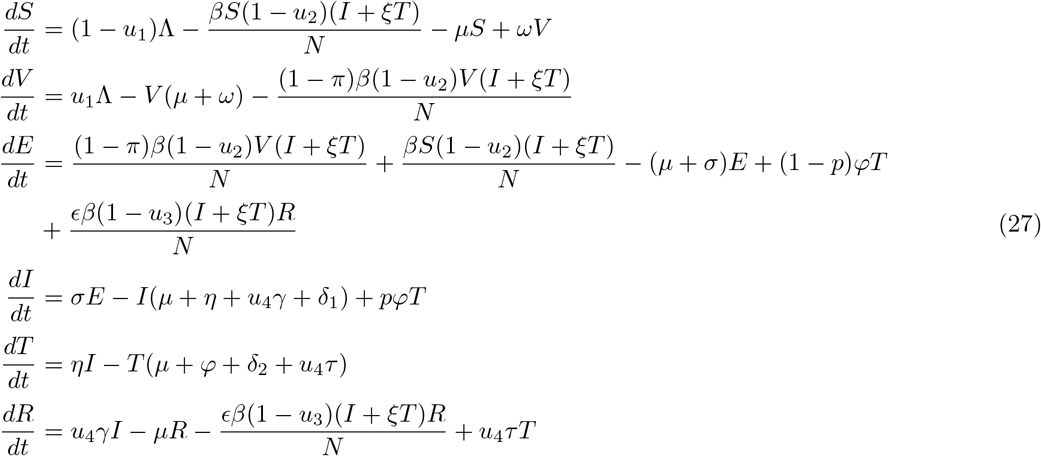

Subject to initial conditions *S*(0) = *S*^0^*, V* (0) = *V*^0^*, E*(0) = *E*^0^*, I*(0) = *I*^0^*, T* (0) = *T*^0^*, R*(0) = *R*^0^ The control *u*_1_*, u*_2_*, u*_3_ and *u*_4_ are bounded, Lesbegue integrable functions. *u*_1_ represents the control through vaccination of a section of the population, *u*_2_ is the control that represents preventive effort against Chlamydia, while *u*_3_ and *u*_4_ represents control parameters accounting for re-infection and treatment control effort respectively.

Objective funtional

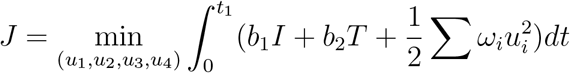

where *t*_1_ is the finaltime. We seek to find an optimal control combinations 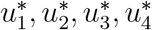, such that

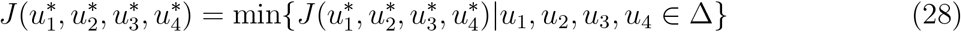

where 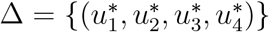 such that 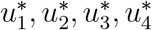 are measurable with 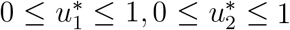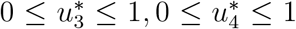 for *t* ∈ [0*, t*_1_] is the control set. Hence, we shall now invetigate the existence of such optimal solution which minimizes the objective functional *J*

**Theorem 5.1**. *Given the objective functional J, defined on the control set* ∆*, and subject to the state system (27) with non-negative initial conditions at t* = 0 *then there exists an optimal control quadruple u*^∗^ = (*u*_1_*, u*_1_*, u*_2_*, u*_3_*, u*_4_) *such that J*(*u*^∗^) = min{*J*(*u*_1_*, u*_2_*, u*_3_*, u*_4_)|*u*_1_*, u*_2_*, u*_3_*, u*_1_ ∈ ∆}

*Similarly, there exists adjoint functions λ*_1_*, λ*_2_*, λ*_3_*, λ*_4_*, λ*_5_*, λ*_6_ *such that*

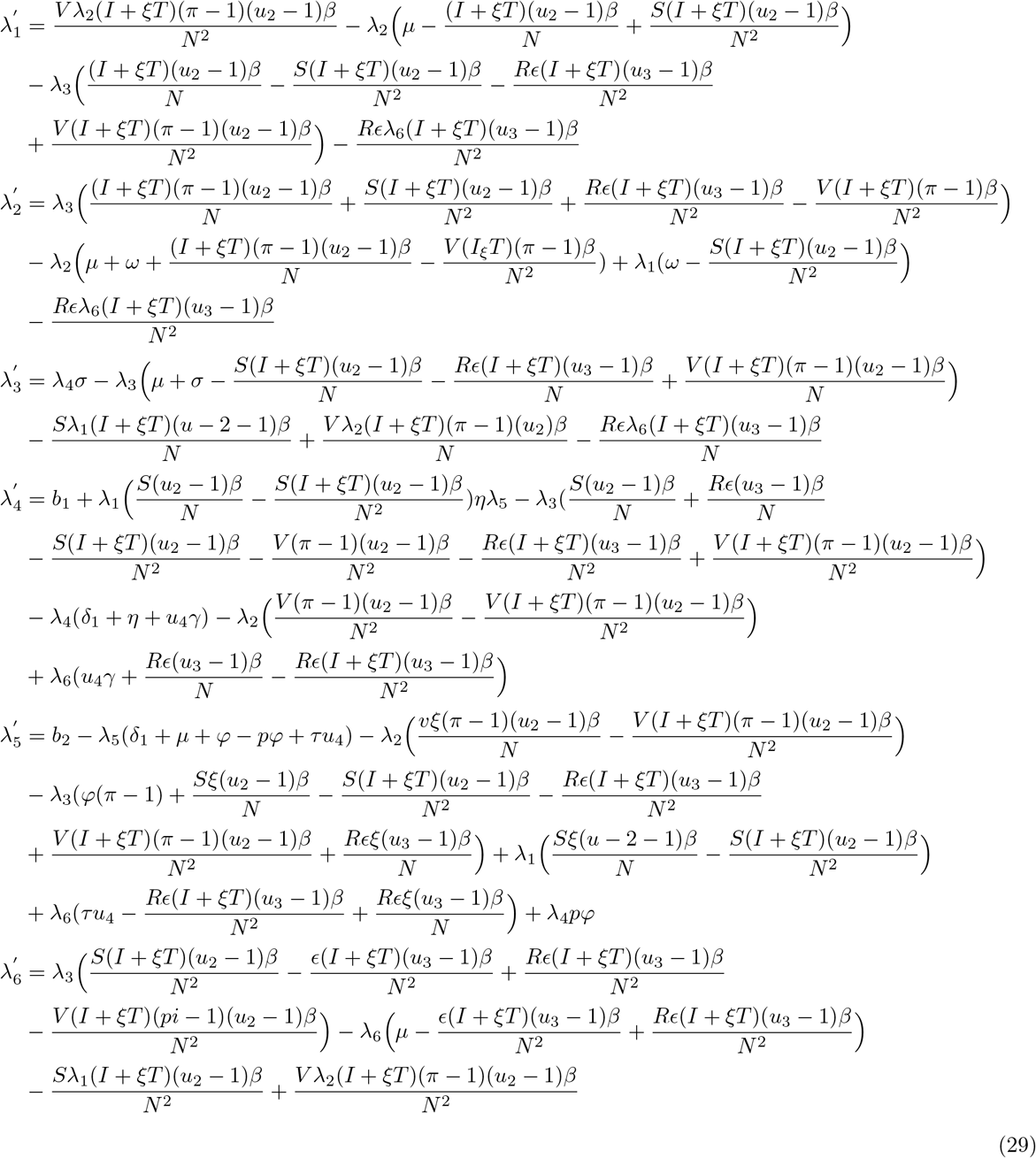

The state function are positive and the controls are lebesgue measurable, therefore we have that *J*(*u_i_*) ≥ 0 for all *u_i_* ∈ *U*. Hence, inf*_u_i*∈*U J*(*u_i_*) exists and is finite. Therefore, a minimizing sequence of controls *u_i_* ∈ *U* exists such that

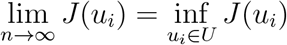

Let *S^n^, V^n^, E^n^, I^n^, T^n^, R^n^* be the associated state trajectories. Since the state sequences are uniformly bounded, we have that the derivatives are also uniformly bounded. As a result, the state sequences are lipschitz continuous with the same constant. Applying the Arzela Ascoli theorem [25], there exits *S*^∗^*, V*^∗^*, E*^∗^*, I*^∗^*, T*^∗^*, R*^∗^ such that on a sub-sequence (*S^n^, V^n^, E^n^, I^n^, T^n^, R^n^*) → *χ*^∗^ uniformly on [0*, t*_1_], where *χ*^∗^ = (*S*^∗^*, V*^∗^*, E*^∗^*, I*^∗^*, T*^∗^*, R*^∗^) since ∥*u_i_*∥_*L*_∞ *< K* for some *K >* 0, it follows that *u_i_* ∈ *L*^2^([0*, t*_1_]), such that on a sub-sequence 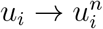 weakly in *L*^2^([0*, t*_1_])*, n* → ∞ Applying the lower semi-continuity of *L*^2^ norm with respect to weak convergence, we have that

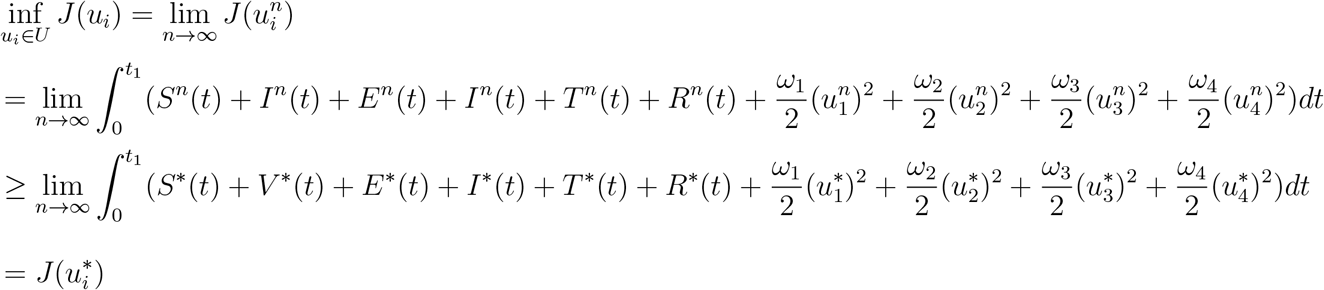

Considering the convergence of the sequence 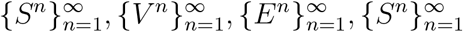, 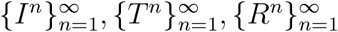 and passing to the limit in the ordinary differential equation system 27, we have *S*^∗^*, V*^∗^*, E*^∗^*, I*^∗^*, T*^∗^*, R*^∗^ are the states corresponding to the control quadruple (*u_i_*).

Hence, *u_i_* is an optimal control quadruple.

From the Pontryagin’s maximum principle, it transforms 27,28,29 into a problem of minimizing a Hamiltonian 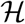, pointwisely with regards to the control functions; *u*_1_*, u*_2_*, u*_3_*, u*_4_

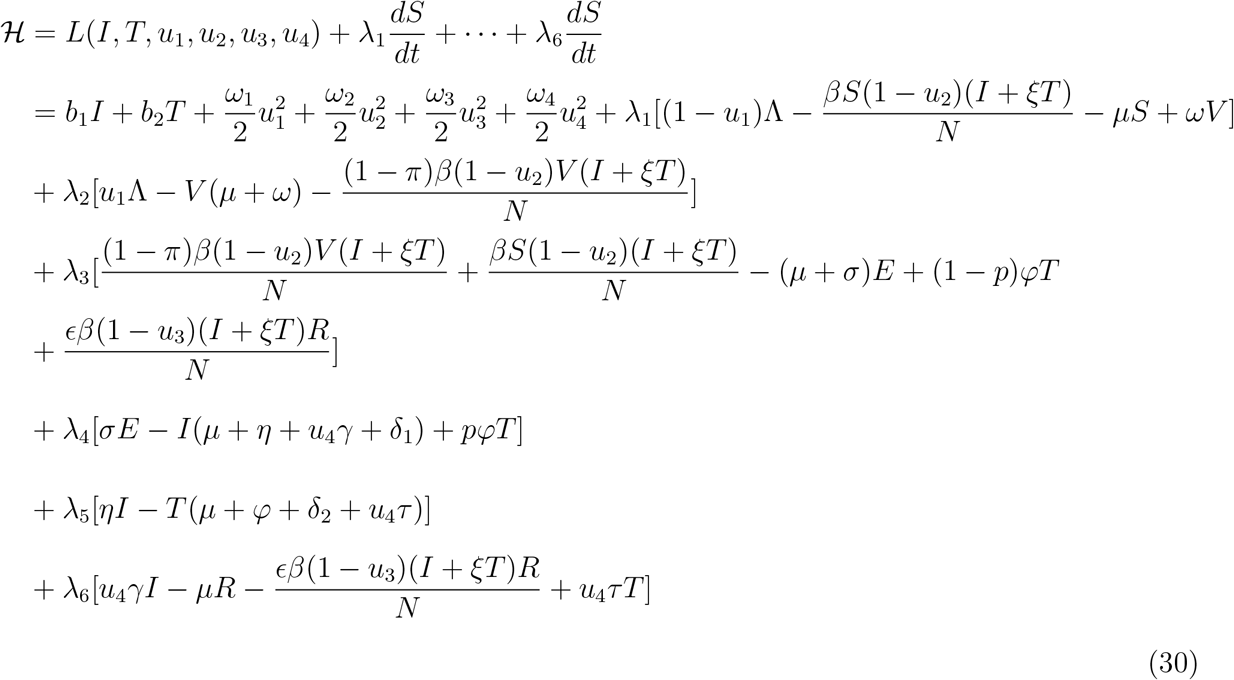

**Theorem 5.2**. *For an optimal control set u*_1_*, u*_2_*, u*_3_*, u*_4_ *that minimizes J over* ∆*, there are adjoint variables λ*_1_*, λ*_2_*, λ*_3_*, λ*_4_*, λ*_5_*, λ*_6_ *satisfying* 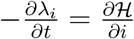 *and transversality conditions*

*λ_i_*(*t_f_*) = 0*, where i* = *S < V < E < I < T < R. Furthermore*

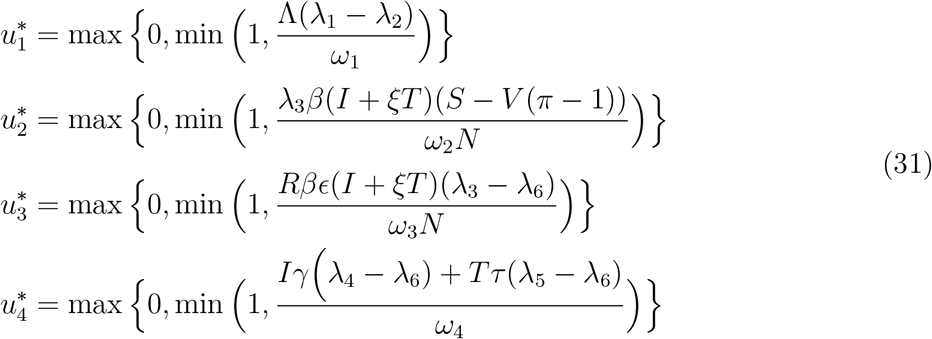

**Proof**

Suppose 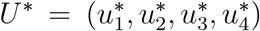 is an optimal control and *S*^∗^*, V*^∗^*, E*^∗^*, I*^∗^*, T*^∗^*, R*^∗^ are the corresponding state solutions.

Applying the Pontrygin’s Maximum Principle, there exists adjoint variables satisfying 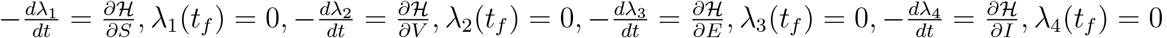, 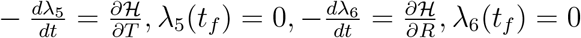 with transversality conditions 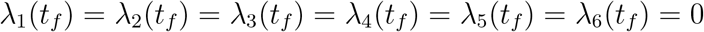

We determine the behaviour of the control by differentiating the Hamiltonian 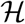 with respect to the controls (*u*_1_*, u*_2_*, u*_3_*, u*_4_) at *t*. On the interior of the control set, where 0 *< u_j_ <* 1 for all *j* = 1, 2, 3, 4, we obtain

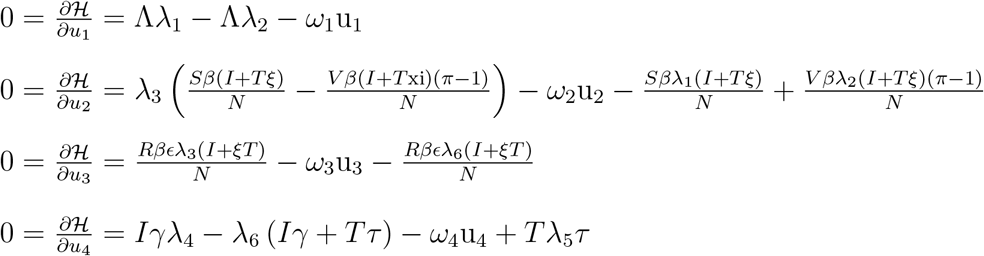

Therefore, we have that

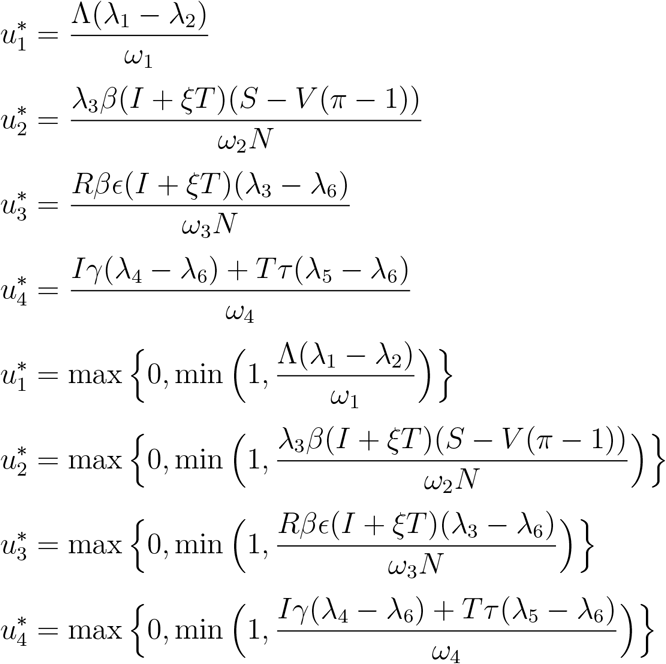

## 6 Sensitivity Analysis of Model Parameters

The sensitivity analysis of the parameters is carried out in this section, which helps to show parameters that have effect on the basic reproduction number. Uncertainties which may result from parameter estimates used in the numerical simulations, a Latin Hyper-cube Sampling (LHS) [30] is applied on the parameters of the model.The Partial Rank Correlation Coefficient (PRCC) between values of the parameters in the response function and the values of the response function was derived from this analysis.

It is observed from the table below that *μ, β* and *σ* varies directly with the reproduction with a significant impact in determining the threshold of the value of the reproduction number, hence an increase in any of these parameters will result to significant increase in the value of the reproduction number. *η* varies inversely with the reproduction number, this implies that an increase in *η* will result to a decrease in the value of 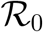. This result is reasonable since *η* is the rate at which the infected becomes treated, hence if increased reduces secondary infection which defines the reproduction number. This same arguement can be extented to *σ* as it describes the movement from exposed to infectious class and hence an increase will also increase the reproduction number.

The effect of the parameters on the other response functions with strong effect on them is depicted in the table in bold font, where when positive varies directly with it and indirectly otherwise.

**Table 2:** PRCC values for the model parameters using the the associated reproduction number 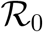, E, I, and T as response functions. Paramters which has significant effect on the dynamics of the model with respect to each of the response functions are shown in bold.

| Parameters | E | I | T | $\mathcal{R}_0$ |
| --- | --- | --- | --- | --- |
| $\mu.$ | **-0.4257** | **-1.4605** | **-0.4359** | **1** |
| $\beta$ | 0.1852 | 0.0630 | 0.0917 | **0.8438** |
| $\varphi$ | **0.7024** | **0.7046** | -0.1639 | 0.1451 |
| $\pi$ | 0.0106 | 0.0988 | 0.0253 | 0.0002102 |
| $\xi$ | 0.0385 | 0.0305 | 0.0228 | 0.2378 |
| $\epsilon$ | 0.0563 | -0.0270 | -0.0773 | 0.0023 |
| $\gamma$ | -0.0262 | -0.3384 | -0.3782 | -0.3146 |
| $\omega$ | -0.1119 | -0.0059 | 0.0104 | -0.2352 |
| $\tau$ | 0.0551 | 0.0627 | -0.0798 | -0.1633 |
| $\delta_1$ | -0.1447 | -0.2811 | -0.3107 | -0.3728 |
| $\delta_2$ | -0.0215 | 0.0195 | -0.0228 | -0.1926 |
| $\sigma$ | -0.1325 | 0.1778 | **0.4119** | **0.8627** |
| $\eta$ | **0.5393** | 0.3614 | **0.8367** | **-0.4903** |
| p | 0.3637 | **0.5782** | **0.8492** | -0.0230 |
| $\phi$ | 0.0652 | -0.0657 | 0.0344 | -0.3054 |

## 7 Numerical Simulations

The optimal control problem is simulated using MATlAB with the following parameter values *b*_1_ = 25*, b*_2_ = 25*, ω*_1_ = 200*, ω*_2_ = 250*, ω*_3_ = 350*, ω*_4_ = 150. It is carried out with the initial conditions *S*(0) = 5000*, V* (0) = 300*, E*(0) = 200*, I*(0) = 200*, T* (0) = 600*, R*(0) = 400. The time interval of the simulation is given as [0,5]. The following strategies are combined to investigate the combination pairs for effective control;

i. Strategy A: *u*_1_ and *u*_3_ = Vaccination and control against re-infection
ii. Strategy B: *u*_1_ and *u*_4_ = Vaccination and treatment effort
iii. Strategy C: *u*_2_ and *u*_3_ = Prevention effort and control against re-infection
iv. Strategy D: *u*_2_ and *u*_4_ = Prevention effort and treatment effort

### 7.0.1 A: combination of optimal controls *u*_1_(*t*) and *u*_3_(*t*)

This strategy considers the vaccination of a fraction of the population and control effort against re-infection and its effect to the population of the infectious and treated class respectively. From figure(2 and 3); it shows that the combination have a positive effect on the population of the afore mentioned classes, which shows a decrease in their population after applying the above stratagies.

**Table 3:** Parameter values

| S/N | Parameter | Symbol | values | Reference |
| --- | --- | --- | --- | --- |
| 1. | Natural death rate | $\mu$ | 0.01245 | estimated. |
| 2. | Recruitment rate | $\Lambda$ | 7233.62 | estimated |
| 3. | waning rate of vaccine | $\omega$ | 0.5 | [6] |
| 4. | Contact rate | $\beta$ | 3.0 | assumed |
| 5. | The rate at which the exposed is infected | $\sigma$ | 0.895 | assumed |
| 6. | Rate of recovery of infectious | $\eta$ | 0.9 | assumed |
| 7. | Death rate due to disease | $\delta_1$ | 0.01 | [6] |
| 8. | Effectiveness of vaccine | $\pi$ | 0.9 | assumed |
| 9. | Fraction of recruited individuals | $\phi$ | 2.0 | [6] |
| 10. | Fraction of treated | p | 0.7 | assumed |
| 10. | Re-infection rate | $\epsilon$ | 0.35 | [6] |
| 11. | Modification parameter | $\xi$ | 0.9 | assumed |
| 15. | Total Population | $N(t)$ | 1000000 | estimated |

**Figure 3:**
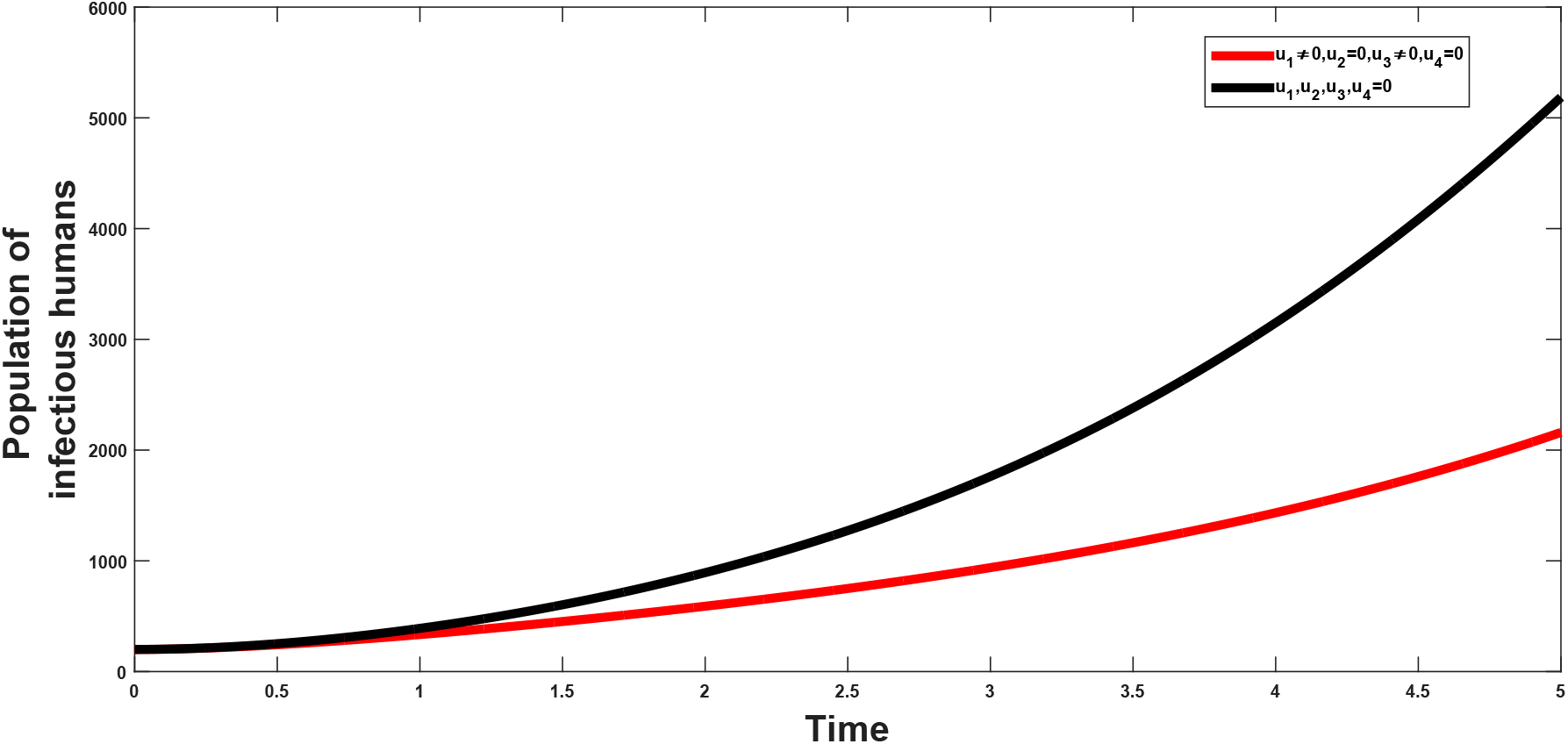
Combined effects of optimal controls *u*_1_ and *u*_3_ on the dynamics of the co-infection optimal control model

**Figure 4:**
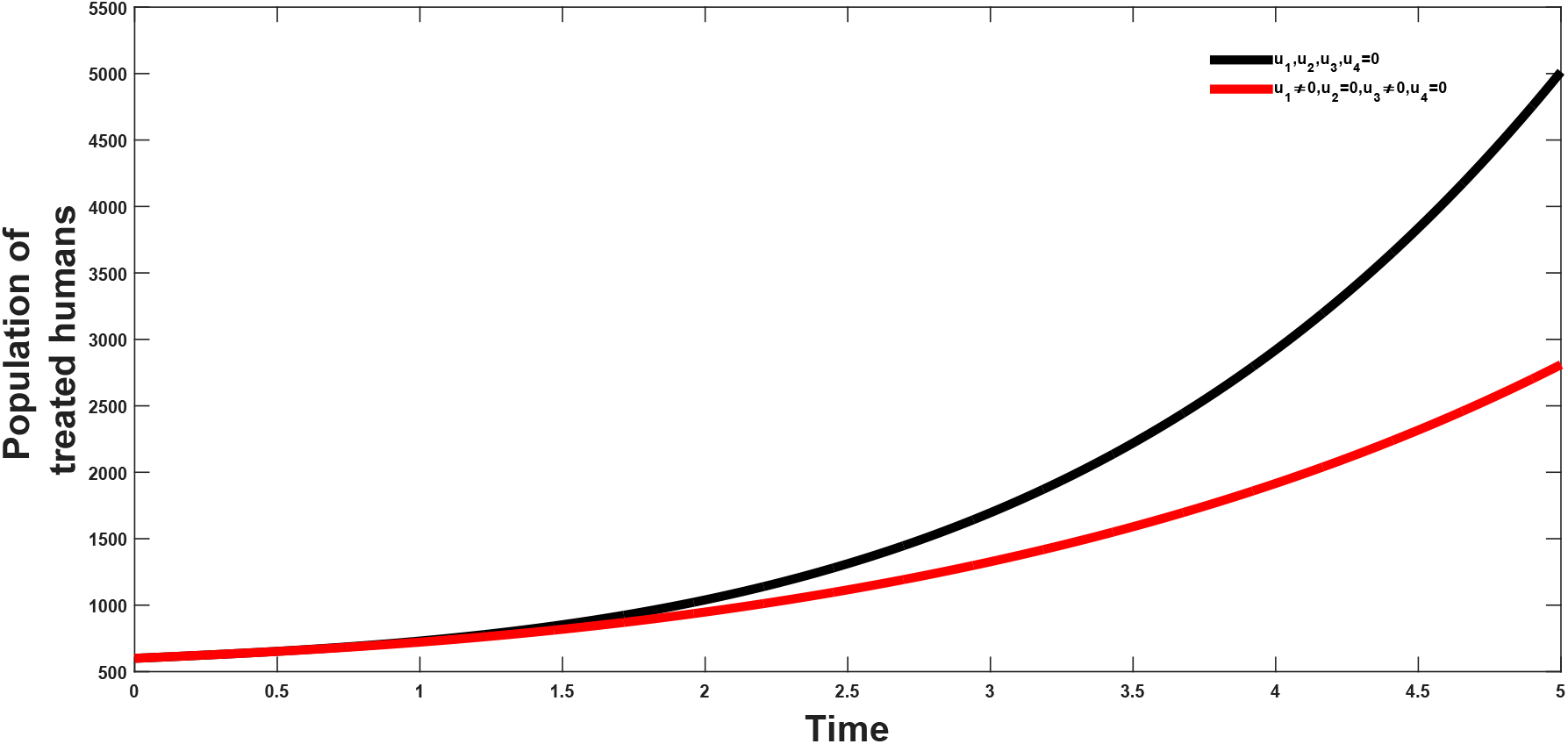
Combined effects of optimal controls *u*_1_ and *u*_3_ on the dynamics infected class

### 7.0.2 B: combination of optimal controls *u*_1_(*t*) and *u*_4_(*t*)

The vaccination and treatment effort are combined to investigate the effect of the combination on both the population of the infectious and the treated population. It is observed that from figure(5 and 6) that this strategy helps in reducing the population of the infectious classes, hence showing that the combination has a positive impact on the population and in the control of the infection.

**Figure 5:**
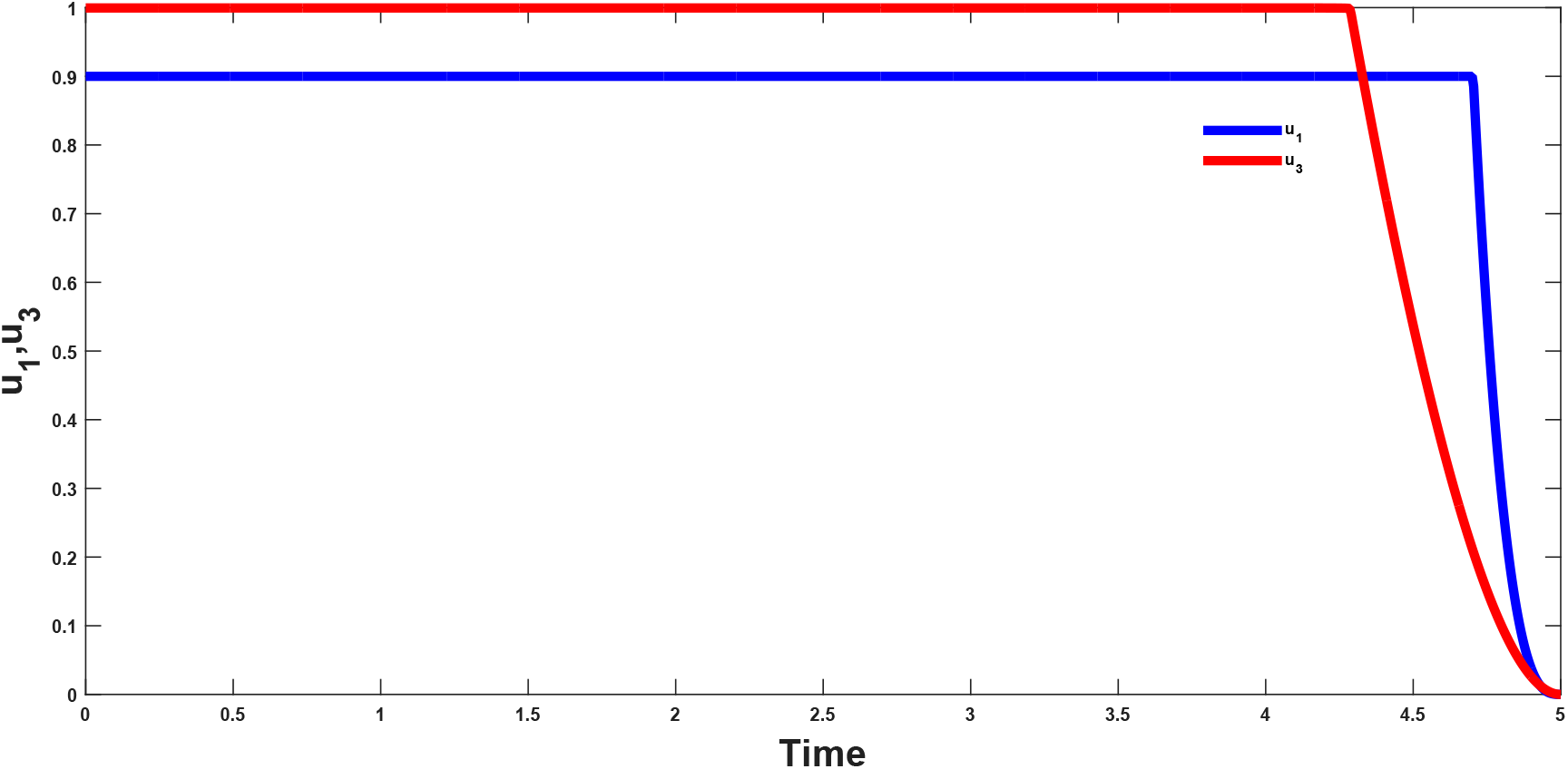
Combined effects of optimal controls *u*_1_ and *u*_3_ on the dynamics of the optimal control model

**Figure 6:**
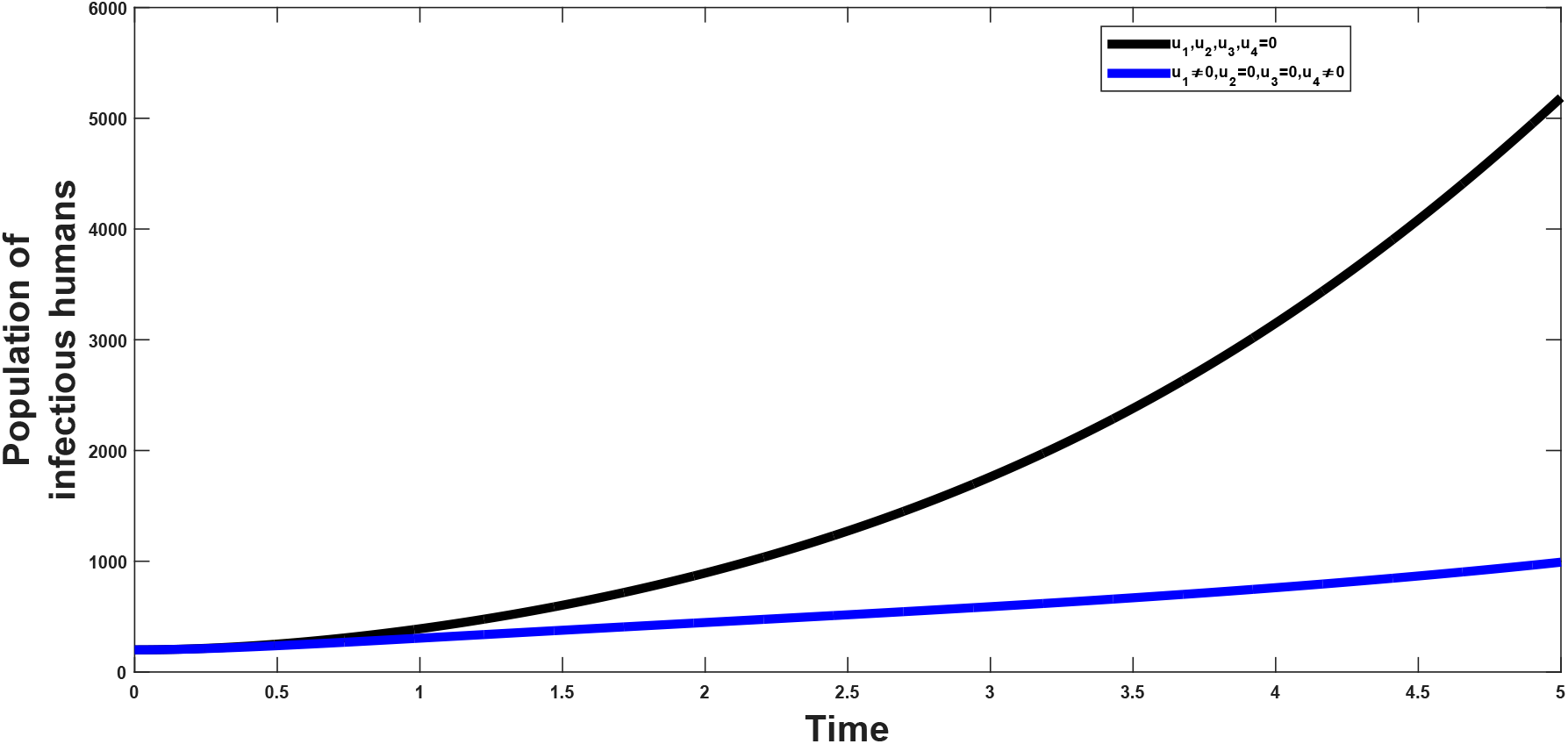
Combined effects of optimal controls *u*_1_ and *u*_4_ on the dynamics infected class

### 7.0.3 C: combination of optimal controls *u*_2_(*t*) and *u*_3_(*t*)

The prevention effort (*u*_2_) with the use of condom and control against re-infection (*u*_3_) is investigated to see their impact on the population. It shows that when these two strategies are combined, the resultant effect on the population is that which reduces the burden of the infection in the populatio. This can be seen from figure(8 and 9).

**Figure 7:**
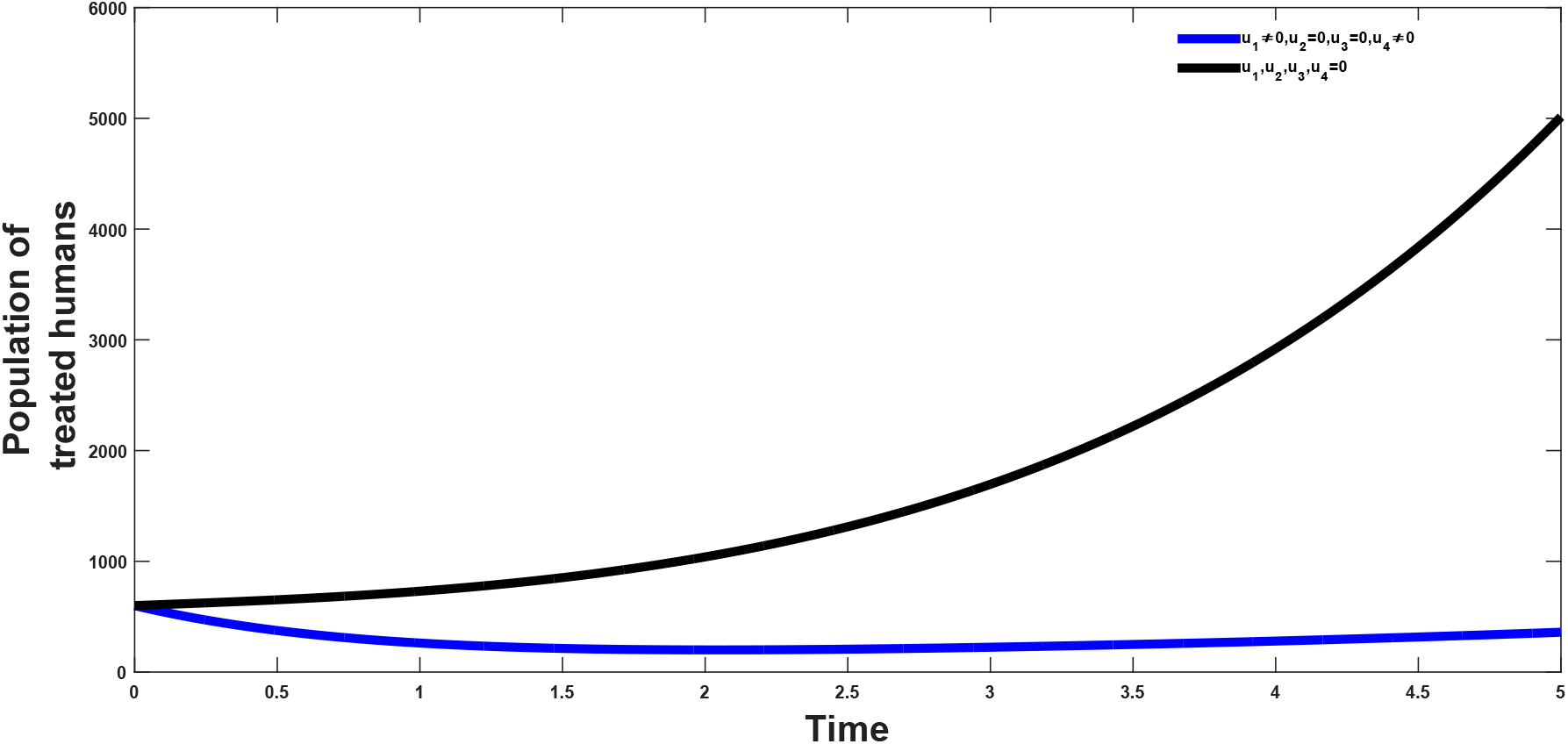
Combined effects of optimal controls *u*_1_ and *u*_4_ on the dynamics treated class

**Figure 8:**
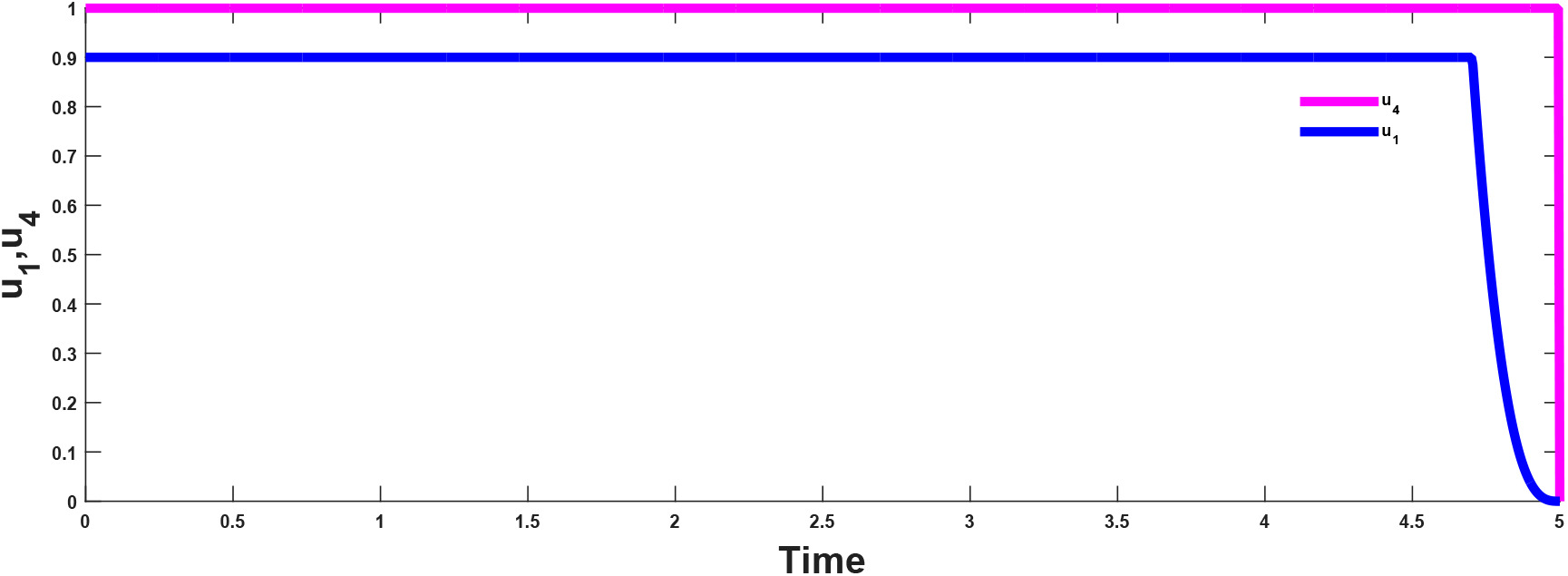
Combined effects of optimal controls *u*_1_ and *u*_4_ on the dynamics of the optimal control model

**Figure 9:**
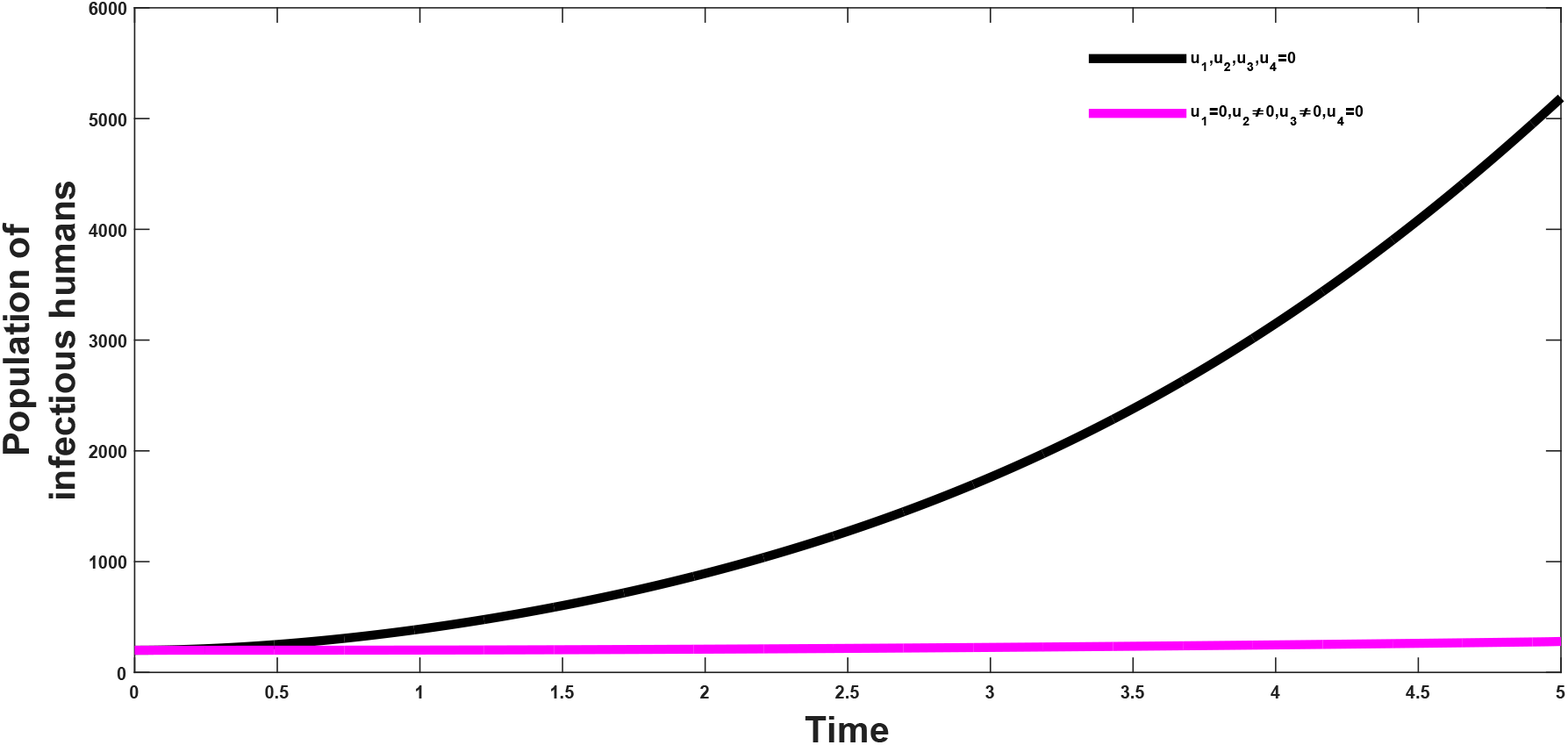
Combined effects of optimal controls *u*_2_ and *u*_3_ on the dynamics of the infected class

**Figure 10:**
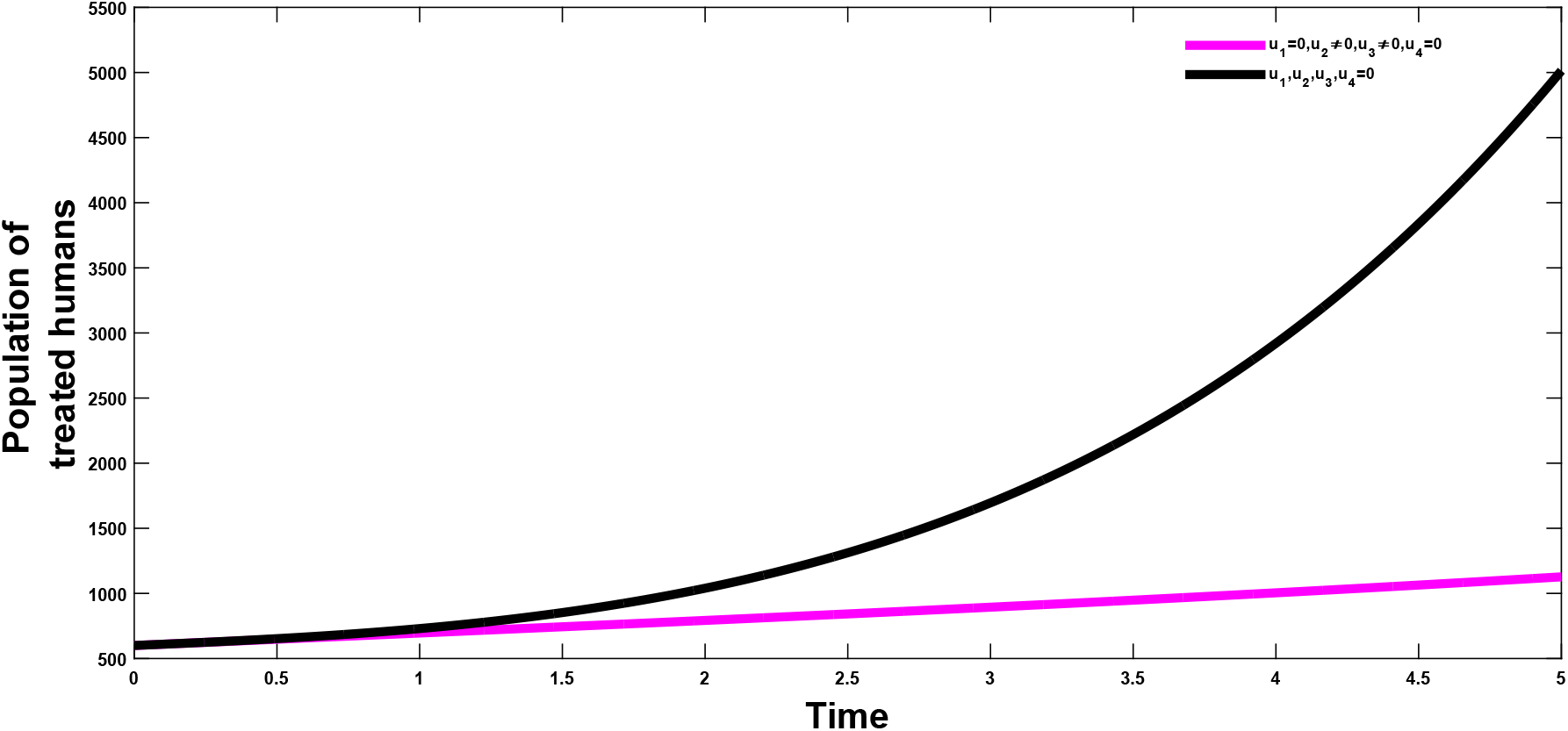
Combined effects of optimal controls *u*_2_ and *u*_3_ on the dynamics of the treated class

**Table 4:**
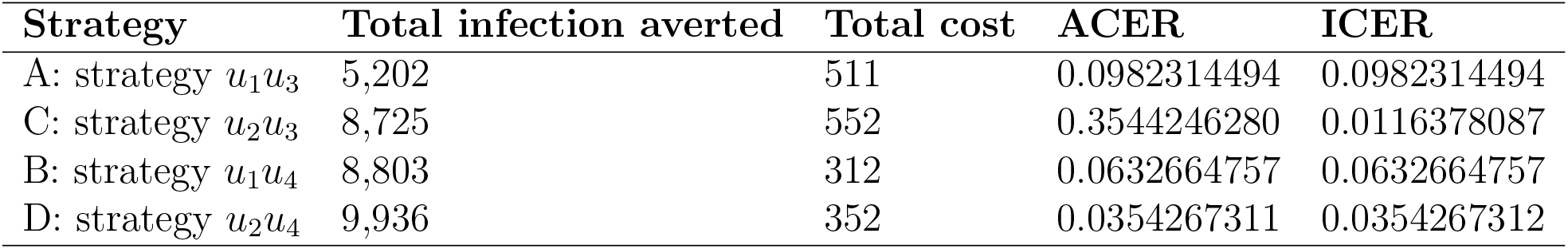
Increasing order of the total infection averted due to the control strategies

### 7.0.4 D: combination of optimal controls *u*_2_(*t*) and *u*_4_(*t*)

When the preventive effort (*u*_2_) and treatment effort (*u*_4_) are combined the plot shows that there will be a reduction of the impact of the infection in the general populace as can be seen from figure(11–12). This strategy highlights the importance of proper treament of patients and prevention effort (proper use of condom) in reducing the burden of the disease in the total population. This, as can be seen in later calculations is the most cost effective strategy and at the same shows a larger disease avertion rate when compared to other strategies.

**Figure 11:**
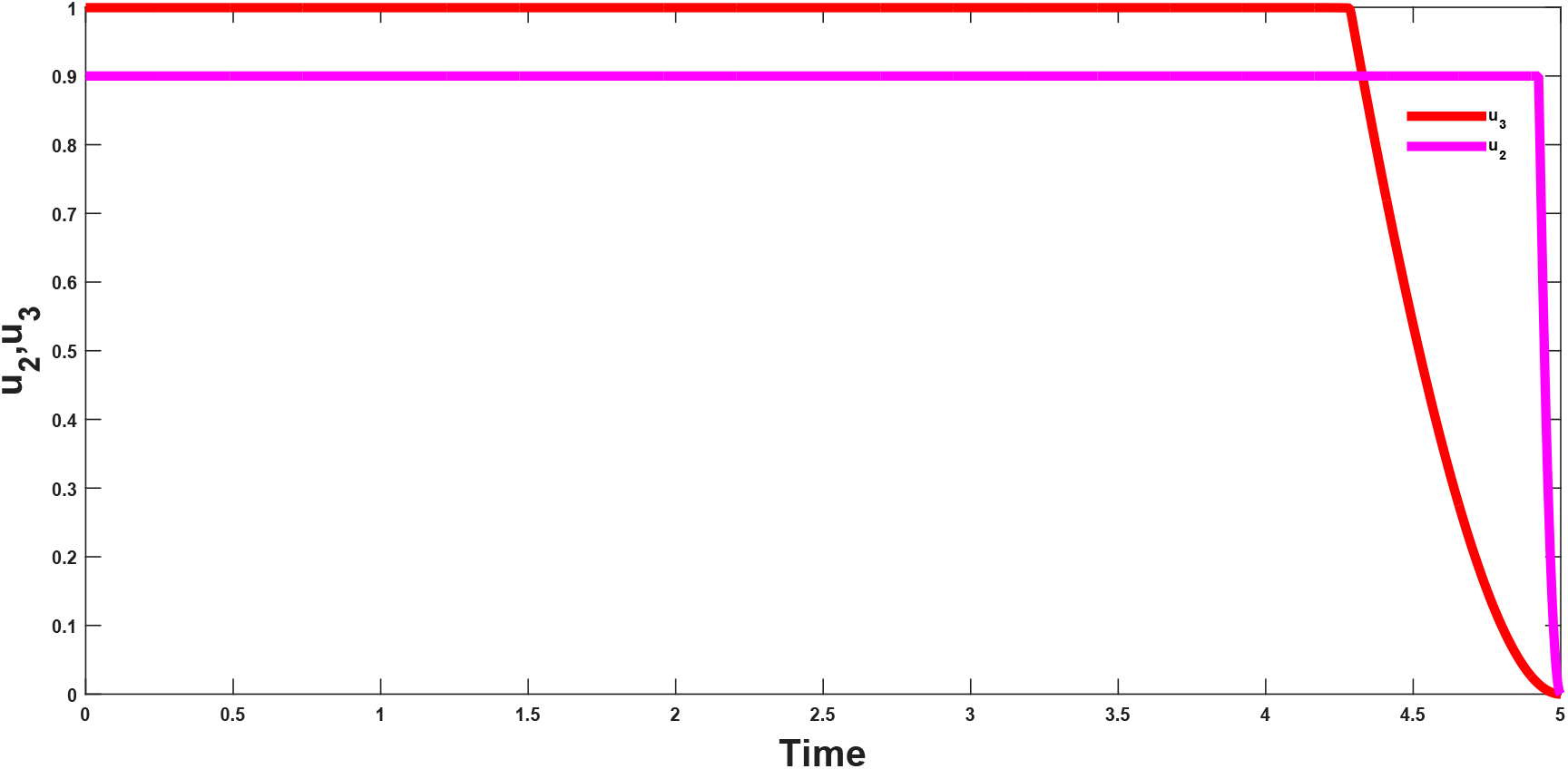
Combined effects of optimal controls *u*_2_ and *u*_3_ on the dynamics of the infected class

**Figure 12:**
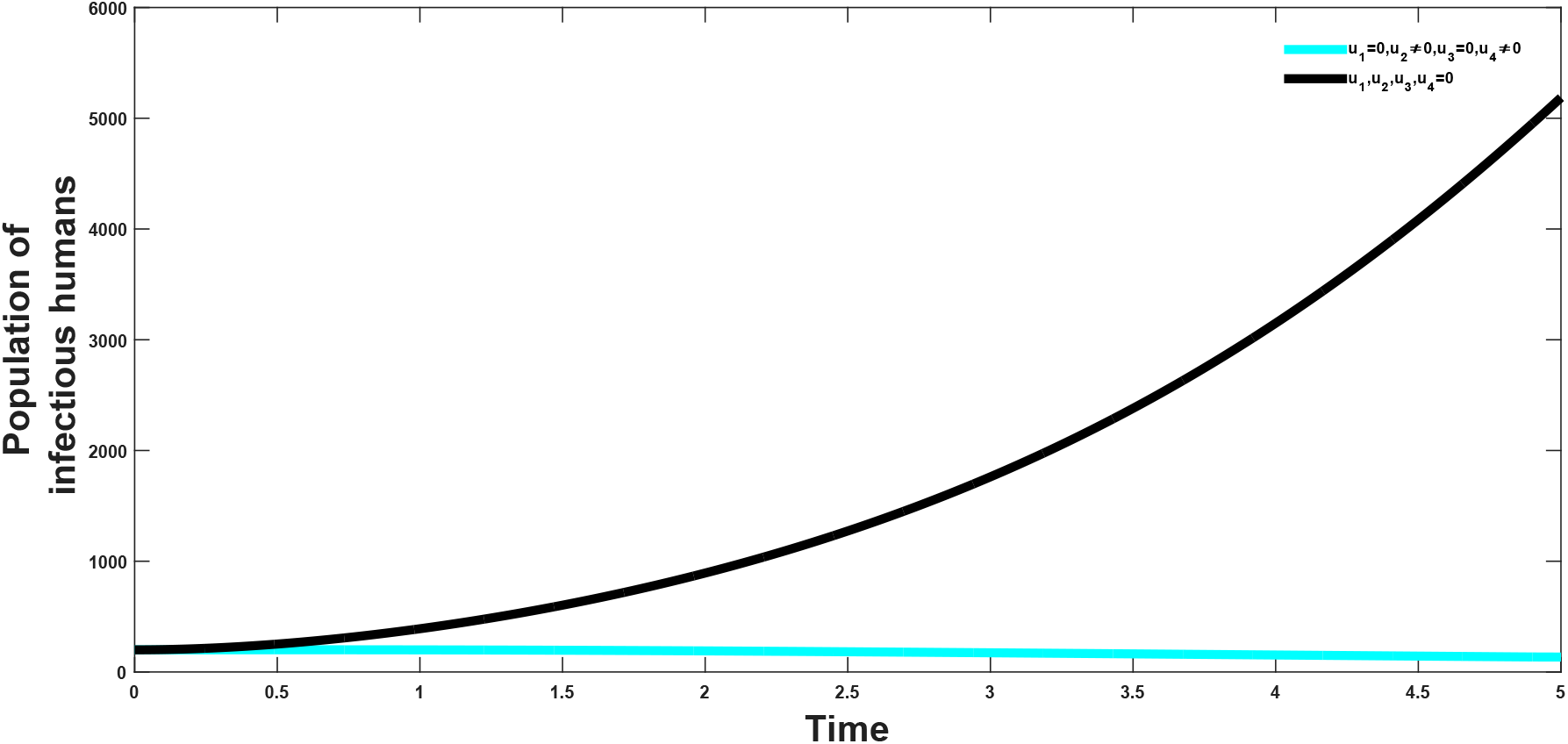
Combined effects of optimal controls *u*_2_ and *u*_4_ on the dynamics of the infected class

### 7.0.5 Cost-Effectiveness Analysis

From the simulations carried out on the optimal control strategies, we then determine the most cost-effective intervention strategies. We use the methods of ACER(age-cost effectivness ratio) and ICER(increamental cost-effectiveness ratio),where

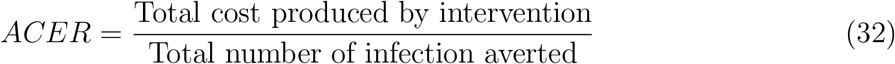

Which focuses on the single intervention strategy and weighing the intervention against its baseline option. Where

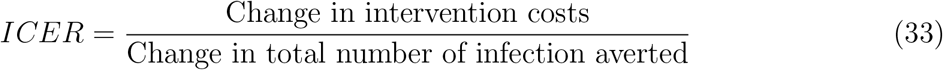

which gives the comparison of differences of two alternative strategies to the change in total number of infection averted by the two strategies.

**Figure 13:**
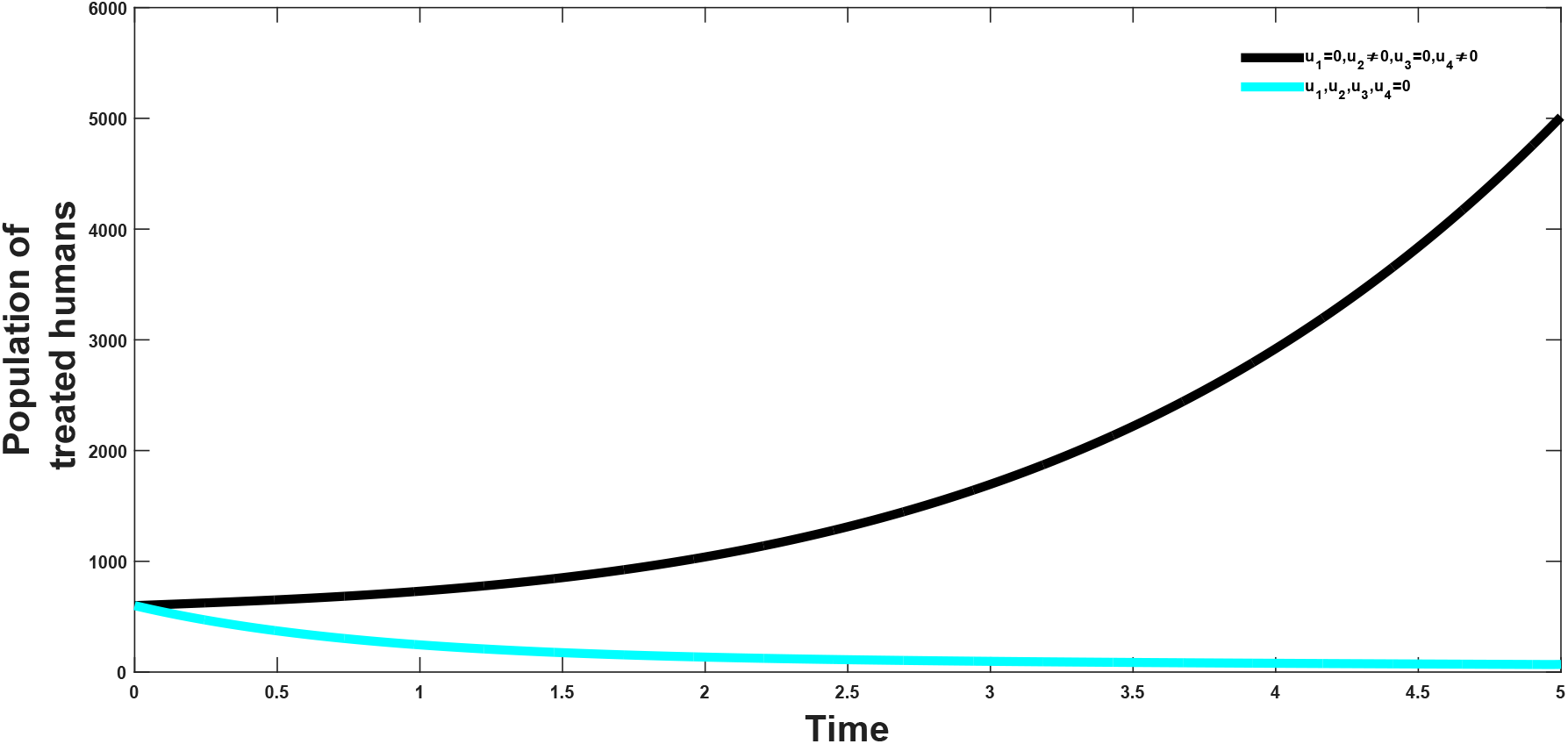
Combined effects of optimal controls *u*_2_ and *u*_4_ on the dynamics of the treated class

**Figure 14:**
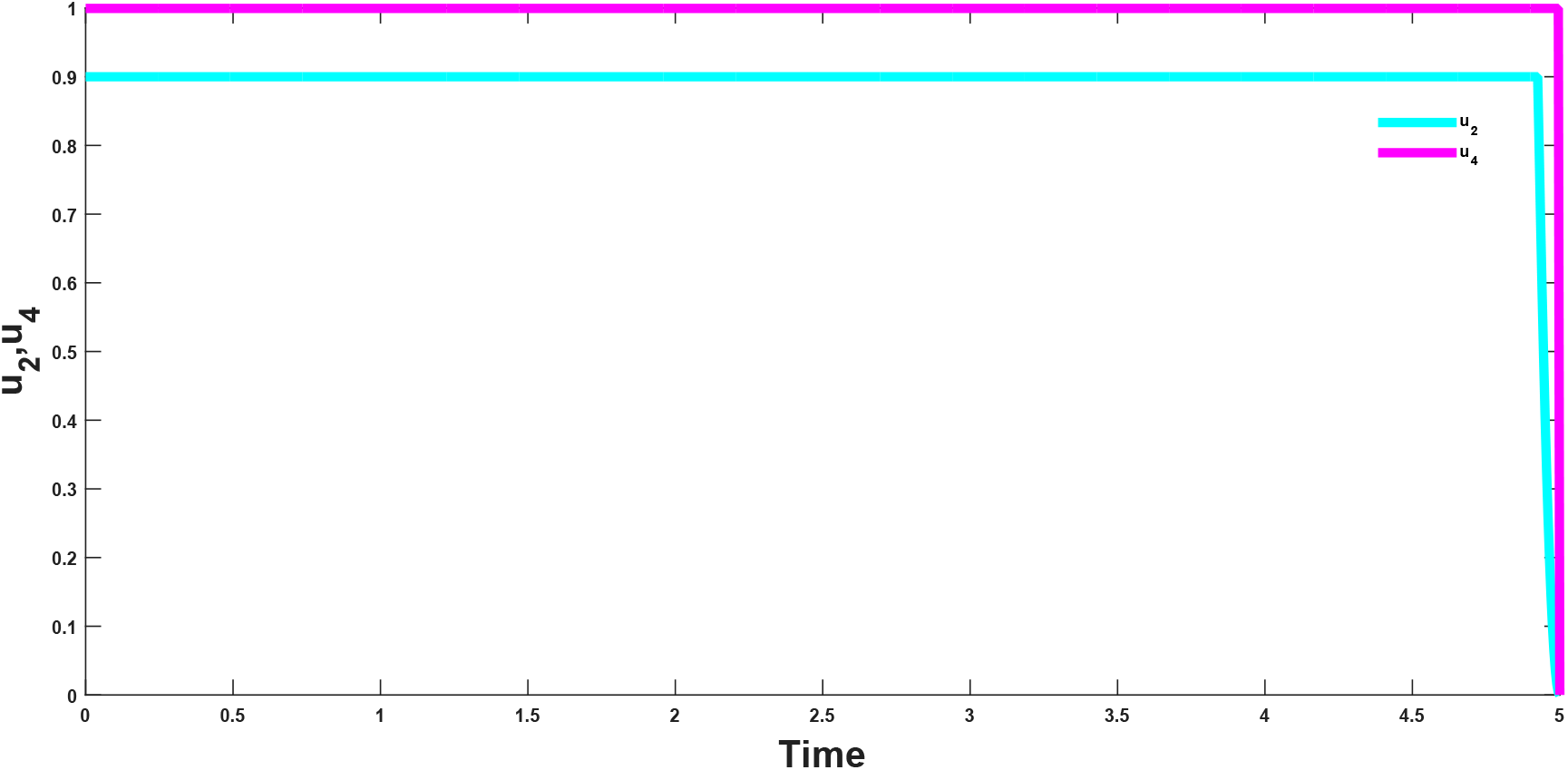
Combined effects of optimal controls *u*_2_ and *u*_4_ on the dynamics of the optimal control model

**Figure 15:**
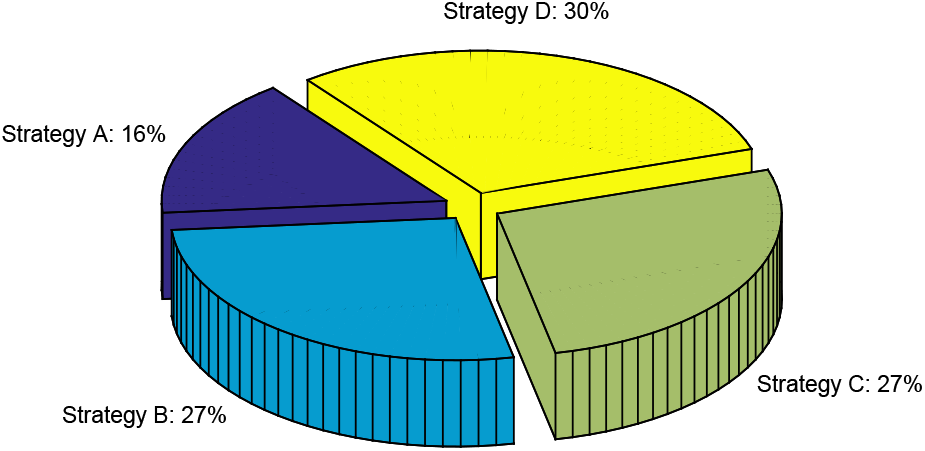
Percentage of total infection averted by differrent strategies

From the table which is in descending order of total infection averted, we calculate the ICER of strategy A and strategy C and the ICER of strategy B and strategy D respectively;

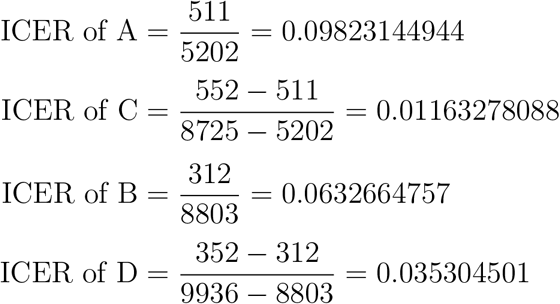

Since C and D has the least cost-effectiveness in comparison to A and B respectively, hence we then compare the ICER of C and D

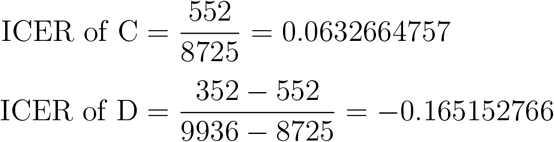

This shows that strategy D (prevention and treatment effort) is the most cost-effective strategy, which also can be seen from the calculation of the AVER. From figure 16, it shows that the universal strategy is the less cost effective strategy

**Table 5:** Increasing order of the total infection averted due to the control strategies

| Strategy | Total infection averted | Total cost | ACER | ICER |
| --- | --- | --- | --- | --- |
| C: strategy $u_2u_3$ | 8,725 | 552 | 0.0632664757 | 0.0632664757 |
| D: strategy $u_2u_4$ | 9,936 | 352 | 0.0354267311 | -0.165152766 |

**Figure 16:**
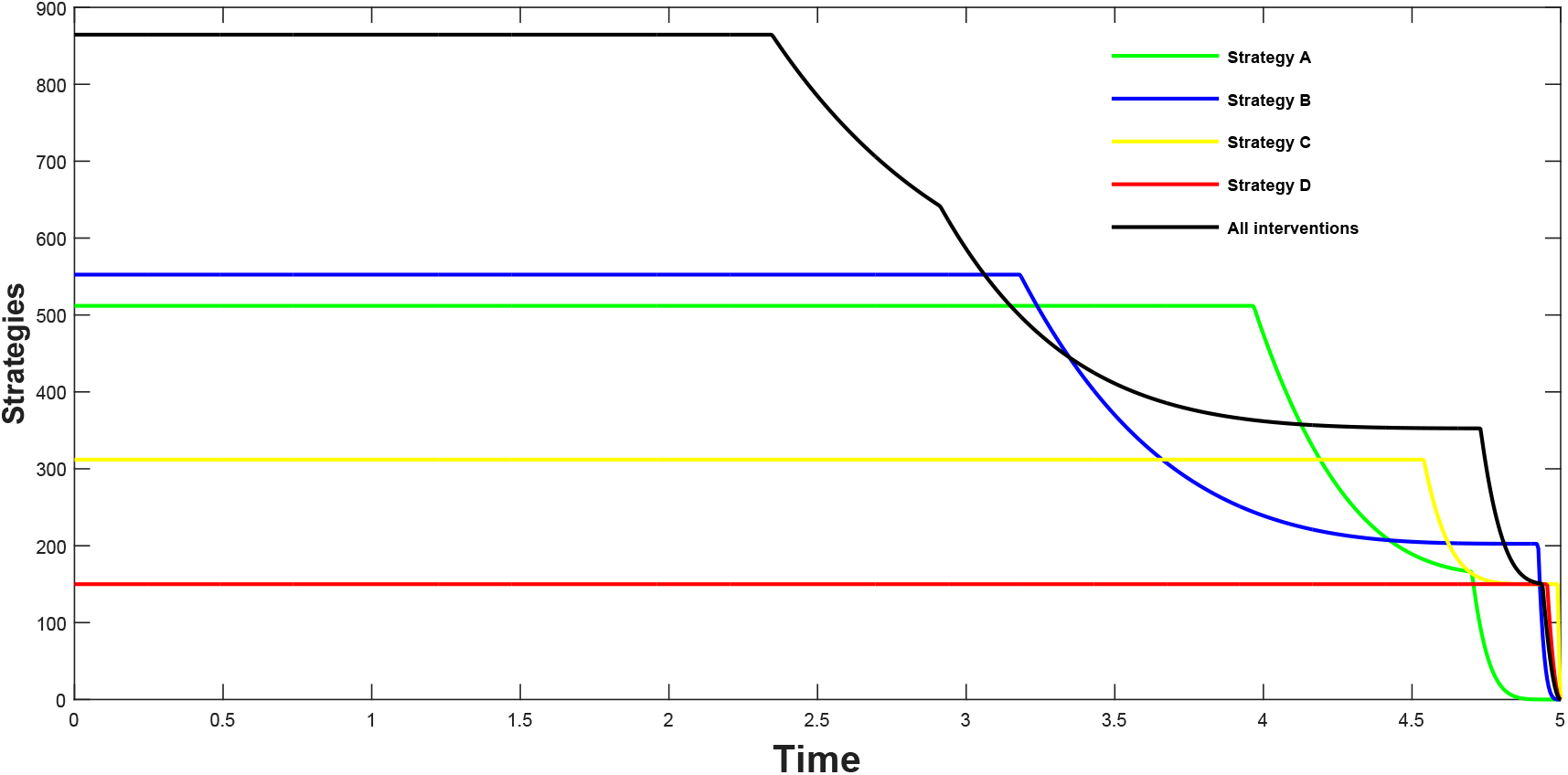
Cost function of the intervention strategies

## 8 Conclusion

In this work we have considered a vaccination model of Chlamydia. The local and global stability of the equlibrium points where established, where it was observed that the model is locally asymptotically stable if the reproduction number is less than unity, and globally stable if a certain threshold value is greater than unity or the re-nfection rate is zero. We also established the necessary conditions for the existence of optimal control and the optimality system for the model was established using the Pontryagin’s Maximum Principle. Sensitivity analysis was also carried out to determine parameters with the most effect in the model, when the associated reproduction numbers, exposed infected, and treated populations were used as response functions. The optimal control of the model shows the effect of different strategies in the transmission dynamicsof the disease and the cost effectiveness of each control pair where the importance of **treatment** and **prevention effort** of an infectious disease in preventing transmission within the population.The reproduction number which helps in determining the rate of spread of the disease, was calculated using the method proposed by van den Driessche and Watmough. The effect of the re-infection rate on the global stability suggests the exhibition of the phenomenon of backward bifurcation of the model. The backward bifurcation of the system was later studied, and it shows that backward bifurcation will occur if the value of the bifurcation parameter ‘a’ is positive.

## Data Availability

Data values and plots are available at demand

